# Analysis of variants in untranslated and promoter regions and breast cancer risk using whole genome sequencing data

**DOI:** 10.1101/2024.07.03.24309763

**Authors:** Naomi Wilcox, Jonathan P. Tyrer, Leila Dorling, Joe Dennis, Marc Naven, Mustapha Abubakar, Thomas U. Ahearn, Irene L. Andrulis, Antonis C. Antoniou, Natalia V. Bogdanova, Stig E. Bojesen, Manjeet K. Bolla, Hiltrud Brauch, Nicola J. Camp, Jenny Chang-Claude, Kamila Czene, Thilo Dörk, D. Gareth Evans, Peter A. Fasching, Jonine D. Figueroa, Henrik Flyger, Eugene J. Gardner, Anna González-Neira, Pascal Guénel, Eric Hahnen, Per Hall, Mikael Hartman, Maartje J. Hooning, Anna Jakubowska, Elza K. Khusnutdinova, Vessela N. Kristensen, Jingmei Li, Annika Lindblom, Artitaya Lophatananon, Arto Mannermaa, Siranoush Manoukian, Roger L. Milne, Rocio Nuñez-Torres, Nadia Obi, Mihalis I. Panayiotidis, Sue K. Park, John R.B. Perry, Muhammad U. Rashid, Emmanouil Saloustros, Elinor J. Sawyer, Marjanka K. Schmidt, Melissa C. Southey, Amanda B. Spurdle, Diana Torres, Qin Wang, Jacques Simard, Soo Hwang Teo, Alison M. Dunning, Peter Devilee, Douglas F. Easton

**Affiliations:** Centre for Cancer Genetic Epidemiology, Department of Public Health and Primary Care, University of Cambridge, Cambridge, UK; Division of Cancer Epidemiology and Genetics, National Cancer Institute, National Institutes of Health, Department of Health and Human Services, Bethesda, MD, USA; Fred A. Litwin Center for Cancer Genetics, Lunenfeld-Tanenbaum Research Institute of Mount Sinai Hospital, Toronto, Ontario, Canada; Department of Molecular Genetics, University of Toronto, Toronto, Ontario, Canada; Department of Radiation Oncology, Hannover Medical School, Hannover, Germany; Gynaecology Research Unit, Hannover Medical School, Hannover, Germany; N.N. Alexandrov Research Institute of Oncology and Medical Radiology, Minsk, Belarus; Copenhagen General Population Study, Herlev and Gentofte Hospital, Copenhagen University Hospital, Herlev, Denmark; Department of Clinical Biochemistry, Herlev and Gentofte Hospital, Copenhagen University Hospital, Herlev, Denmark; Faculty of Health and Medical Sciences, University of Copenhagen, Copenhagen, Denmark; Dr. Margarete Fischer-Bosch-Institute of Clinical Pharmacology, Stuttgart, Germany; iFIT-Cluster of Excellence, University of Tübingen, Tübingen, Germany; German Cancer Consortium (DKTK) and German Cancer Research Center (DKFZ), Partner Site Tübingen, Tübingen, Germany; Department of Internal Medicine and Huntsman Cancer Institute, University of Utah, Salt Lake City, UT, USA; Division of Cancer Epidemiology, German Cancer Research Center (DKFZ), Heidelberg, Germany; Cancer Epidemiology Group, University Cancer Center Hamburg (UCCH), University Medical Center Hamburg-Eppendorf, Hamburg, Germany; Department of Medical Epidemiology and Biostatistics, Karolinska Institutet, Stockholm, Sweden; Division of Evolution and Genomic Sciences, School of Biological Sciences, Faculty of Biology, Medicine and Health, University of Manchester, Manchester Academic Health Science Centre, Manchester, UK; North West Genomics Laboratory Hub, Manchester Centre for Genomic Medicine, St Mary’s Hospital, Manchester University NHS Foundation Trust, Manchester Academic Health Science Centre, Manchester, UK; Department of Gynecology and Obstetrics, Comprehensive Cancer Center Erlangen-EMN, Friedrich-Alexander University Erlangen-Nuremberg, University Hospital Erlangen, Erlangen, Germany; Usher Institute of Population Health Sciences and Informatics, The University of Edinburgh, Edinburgh, UK; Cancer Research UK Edinburgh Centre, The University of Edinburgh, Edinburgh, UK; Department of Breast Surgery, Herlev and Gentofte Hospital, Copenhagen University Hospital, Herlev, Denmark; MRC Epidemiology Unit, Wellcome-MRC Institute of Metabolic Science, University of Cambridge, Cambridge, UK; Human Genotyping Unit-CeGen, Spanish National Cancer Research Centre (CNIO), Madrid, Spain; Team ‘Exposome and Heredity’, CESP, Gustave Roussy, INSERM, University Paris-Saclay, UVSQ, Villejuif, France; Center for Familial Breast and Ovarian Cancer, Faculty of Medicine and University Hospital Cologne, University of Cologne, Cologne, Germany; Center for Integrated Oncology (CIO), Faculty of Medicine and University Hospital Cologne, University of Cologne, Cologne, Germany; Department of Oncology, Södersjukhuset, Stockholm, Sweden; Saw Swee Hock School of Public Health, National University of Singapore and National University Health System, Singapore City, Singapore; Department of Surgery, National University Health System, Singapore City, Singapore; Department of Pathology, Yong Loo Lin School of Medicine, National University of Singapore, Singapore City, Singapore; Department of Medical Oncology, Erasmus MC Cancer Institute, Rotterdam, the Netherlands; Independent Laboratory of Molecular Biology and Genetic Diagnostics, Pomeranian Medical University, Szczecin, Poland; International Hereditary Cancer Center, Department of Genetics and Pathology, Pomeranian Medical University in Szczecin, Szczecin, Poland; Institute of Biochemistry and Genetics of the Ufa Federal Research Centre of the Russian Academy of Sciences, Ufa, Russia; Department of Genetics and Fundamental Medicine, Bashkir State University, Ufa, Russia; Department of Medical Genetics, Oslo University Hospital and University of Oslo, Oslo, Norway; Institute of Clinical Medicine, Faculty of Medicine, University of Oslo, Oslo, Norway; Human Genetics Division, Genome Institute of Singapore, Agency for Science, Technology and Research (A*STAR), Singapore City, Singapore; Department of Molecular Medicine and Surgery, Karolinska Institutet, Stockholm, Sweden; Department of Clinical Genetics, Karolinska University Hospital, Stockholm, Sweden; Division of Population Health, Health Services Research and Primary Care, School of Health Sciences, Faculty of Biology, Medicine and Health, The University of Manchester, Manchester, UK; Translational Cancer Research Area, University of Eastern Finland, Kuopio, Finland; Institute of Clinical Medicine, Pathology and Forensic Medicine, University of Eastern Finland, Kuopio, Finland; Biobank of Eastern Finland, Kuopio University Hospital, Kuopio, Finland; Unit of Medical Genetics, Department of Medical Oncology and Hematology, Fondazione IRCCS Istituto Nazionale dei Tumori di Milano, Milan, Italy; Cancer Epidemiology Division, Cancer Council Victoria, Melbourne, Victoria, Australia; Centre for Epidemiology and Biostatistics, Melbourne School of Population and Global Health, The University of Melbourne, Melbourne, Victoria, Australia; Precision Medicine, School of Clinical Sciences at Monash Health, Monash University, Clayton, Victoria, Australia; Human Cancer Genetics Programme, Spanish National Cancer Research Centre (CNIO), Madrid, Spain; Institute for Occupational and Maritime Medicine, University Medical Center Hamburg-Eppendorf, Hamburg, Germany; Institute for Medical Biometry and Epidemiology, University Medical Center Hamburg-Eppendorf, Hamburg, Germany; Department of Cancer Genetics, Therapeutics and Ultrastructural Pathology, The Cyprus Institute of Neurology & Genetics, Nicosia, Cyprus; Department of Preventive Medicine, Seoul National University College of Medicine, Seoul, Korea; Integrated Major in Innovative Medical Science, Seoul National University College of Medicine, Seoul, Korea; Cancer Research Institute, Seoul National University, Seoul, Korea; Metabolic Research Laboratory, Wellcome-MRC Institute of Metabolic Science, University of Cambridge, Cambridge, UK; Molecular Genetics of Breast Cancer, German Cancer Research Center (DKFZ), Heidelberg, Germany; Department of Basic Sciences, Shaukat Khanum Memorial Cancer Hospital and Research Centre (SKMCH & RC), Lahore, Pakistan; Department of Oncology, University Hospital of Larissa, Larissa, Greece; School of Cancer & Pharmaceutical Sciences, Comprehensive Cancer Centre, Guy’s Campus, King’s College London, London, UK; Division of Molecular Pathology, The Netherlands Cancer Institute, Amsterdam, the Netherlands; Division of Psychosocial Research and Epidemiology, The Netherlands Cancer Institute - Antoni van Leeuwenhoek hospital, Amsterdam, the Netherlands; Department of Clinical Genetics, Leiden University Medical Center, Leiden, the Netherlands; Department of Clinical Pathology, The University of Melbourne, Melbourne, Victoria, Australia; Population Health Program, QIMR Berghofer Medical Research Institute, Brisbane, Queensland, Australia; Institute of Human Genetics, Pontificia Universidad Javeriana, Bogota, Colombia; Genomics Center, Centre Hospitalier Universitaire de Québec – Université Laval Research Center, Québec City, Québec, Canada; Breast Cancer Research Programme, Cancer Research Malaysia, Subang Jaya, Selangor, Malaysia; Department of Surgery, Faculty of Medicine, University of Malaya, UM Cancer Research Institute, Kuala Lumpur, Malaysia; Centre for Cancer Genetic Epidemiology, Department of Oncology, University of Cambridge, Cambridge, UK; Department of Pathology, Leiden University Medical Center, Leiden, the Netherlands; Department of Human Genetics, Leiden University Medical Center, Leiden, the Netherlands

## Abstract

Recent exome-wide association studies have explored the role of coding variants in breast cancer risk, highlighting the role of rare variants in multiple genes including *BRCA1, BRCA2, CHEK2, ATM* and *PALB2*, as well as new susceptibility genes e.g., *MAP3K1*. These genes, however, explain a small proportion of the missing heritability of the disease. Much of the missing heritability likely lies in the non-coding genome. We evaluated the role of rare variants in the 5’ and 3’ untranslated regions (UTRs) of 18,676 genes, and 35,201 putative promoter regions, using whole-genome sequencing data from UK Biobank on 8,001 women with breast cancer and 92,534 women without breast cancer. Burden tests and SKAT-O tests were performed in UTR and promoter regions. For UTR regions of 35 putative breast cancer susceptibility genes, we additionally performed a meta-analysis with a large breast cancer case-control dataset. Associations for 8 regions at P<0.0001 were identified, including several with known roles in tumorigenesis. The strongest evidence of association was for variants in the 5’UTR of *CDK5R1* (P=8.5×10^−7^). These results highlight the potential role of non-coding regulatory regions in breast cancer susceptibility.

## Introduction

Genetic susceptibility to breast cancer is known to be conferred by common variants, identified through genome-wide association studies (GWAS), together with rare coding variants conferring higher disease risks. Protein-truncating variants (PTVs) and rare missense variants identified through linkage and targeting sequencing studies in some genes have been well established, including *ATM, BARD1, BRCA1, BRCA2, CHEK2, RAD51C, RAD51D, PALB2* and *TP53*^1^. Recently, whole exome sequencing (WES) analysis identified additional associations for PTVs and rare missense variants in *MAP3K1, LZTR1, ATRIP* and *CDKN2A*^2^. However, these common and rare variants, in aggregate, only explain ~50% of the familial aggregation of the disease. Much of the “missing” heritability is likely to be due to variants in the non-coding genome^2^. Here we explore the association of variants in non-coding untranslated regions (UTRs) and promoter regions with breast cancer risk.

UTRs are regions upstream (5’ UTR) and downstream (3’ UTR) of the coding sequence of a gene that are transcribed but not translated into protein^3^, while promoters are untranscribed regions upstream of genes where proteins bind to initiate transcription^4^. Variants in these regions do not affect the protein-coding sequence but may regulate gene expression. We used whole genome sequencing (WGS) data from the UK Biobank (UKB), to assess the role of rare variants in promoter and UTR regions in all genes. After quality control (QC; see methods), this dataset comprised 8,001 women with breast cancer and 92,534 women without breast cancer (Supplementary Table 1). For 35 putative breast cancer susceptibility genes, we incorporated additional data from 51,494 women with breast cancer and 43,884 women without breast cancer from the BRIDGES dataset^1^.

For promoter regions, we considered all rare variants (minor allele frequency<0.001) in promoter regions defined by Ensembl BioMart^5^ excluding variants falling in coding sequences. For UTR regions, we considered all rare variants annotated as a UTR variant according to Ensembl Variant Effect Predictor (VEP)^6^.

We conducted burden tests for all promoter regions or UTR regions in which there was at least one carrier of a variant in either cases or controls (35,201 promoter regions, 16,381 3’ UTRs and 17,185 5’ UTRs). These tests, in which variants are collapsed together, can be more powerful than single-variant tests if variants have similar effect sizes^7^. We used logistic regression with all rare variants grouped together and further improved power by incorporating data on family history of breast cancer, as described elsewhere^2^. Since the assumption of similar effect sizes may break down for these regulatory regions^8^, for burden analyses with P<0.05, we conducted robust SKAT-O tests, which allow for variants to have different effect sizes and directions of association. We denote P-values from the burden test as P_B_, P-values from the robust SKAT-O test as P_S,_ and P-values from the meta-analysis of burden results for the 35 putative breast cancer susceptibility genes as P_BM_.

## Results

### 5’ UTRs

23 5’ UTR regions were associated at P_B_<0.001 (Figures 1 and 2; Supplementary Table 2), slightly more than the number (17) expected by chance. 21 of these corresponded to an increased risk, compared with ~8.5 which would be expected by chance. SKAT-O robust was tested on 861 5’ UTR regions with P_B_<0.05; 14 of these had P_S_<0.001 (Figure 3, Supplementary Table 3). Associations with P_B_ <1×10^−4^ or P_S_ <1×10^−4^ were observed for: *CDK5R1* (P_B_ =8.5×10^−7^, P_S_ =7.2×10^−7^), *MVB12A* (P_B_ =6.6×10^−5^, P_S_ =1.6×10^−4^) and *SYNE1* (P_B_ =8.0×10^−5^, 0.014). None of the five most clearly established breast cancer risk genes had P_B_ or P_S_ <0.001 (Supplementary Table 4).

**Figure 1:**
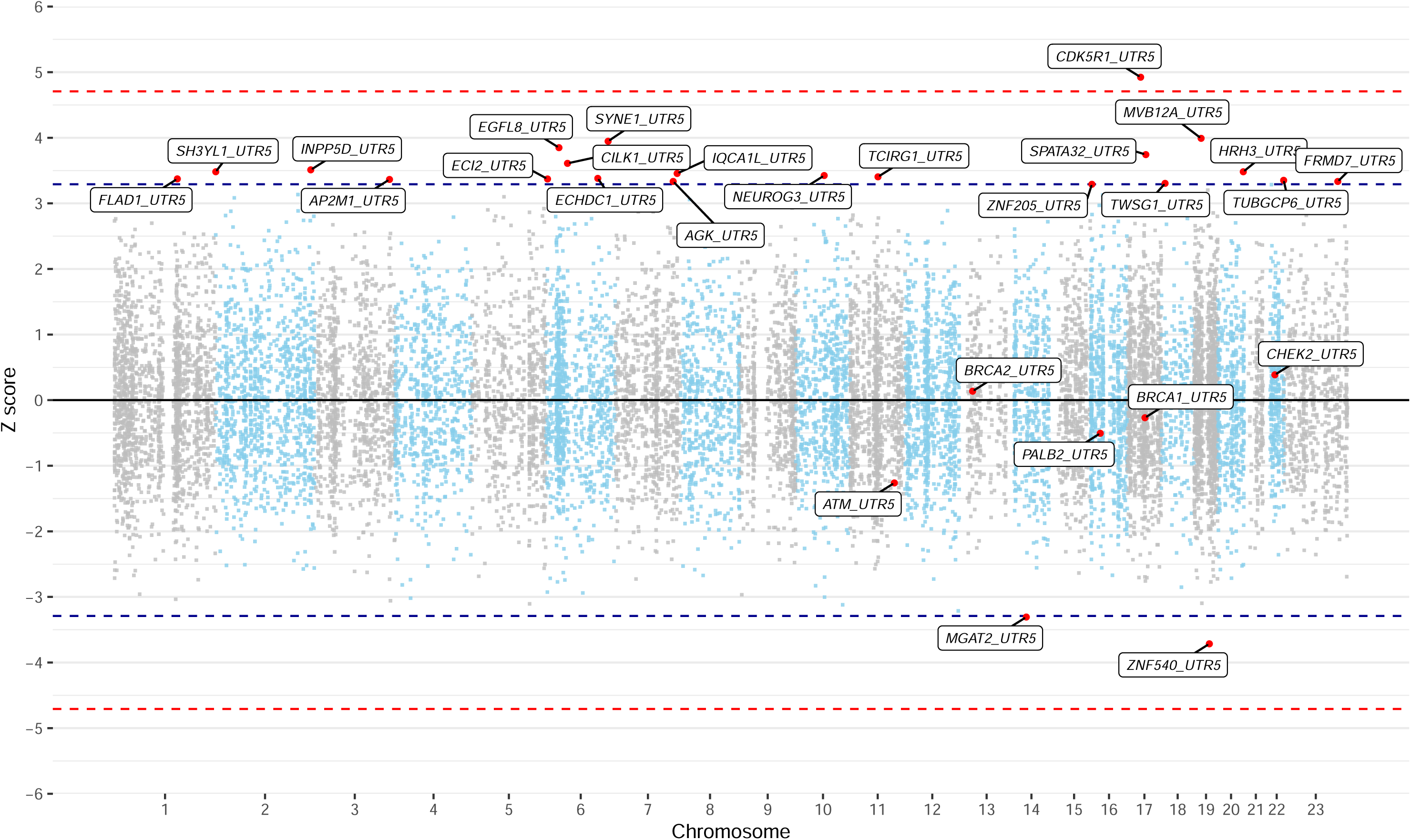
Manhattan Plot of Z-scores from assessing the association between rare variant carriers in 5’ UTR regions and breast cancer risk with the burden test. The x-axis is the chromosomal position, and the y-axis is the Z-score from testing H0 of no association. The blue lines correspond to Z=±3.29, P=0.001, and the red lines correspond to Z=±4.71, P=2.5×10^−6^. All labelled genes are those with P<0.001. P-values are unadjusted for multiple testing.

**Figure 2:**
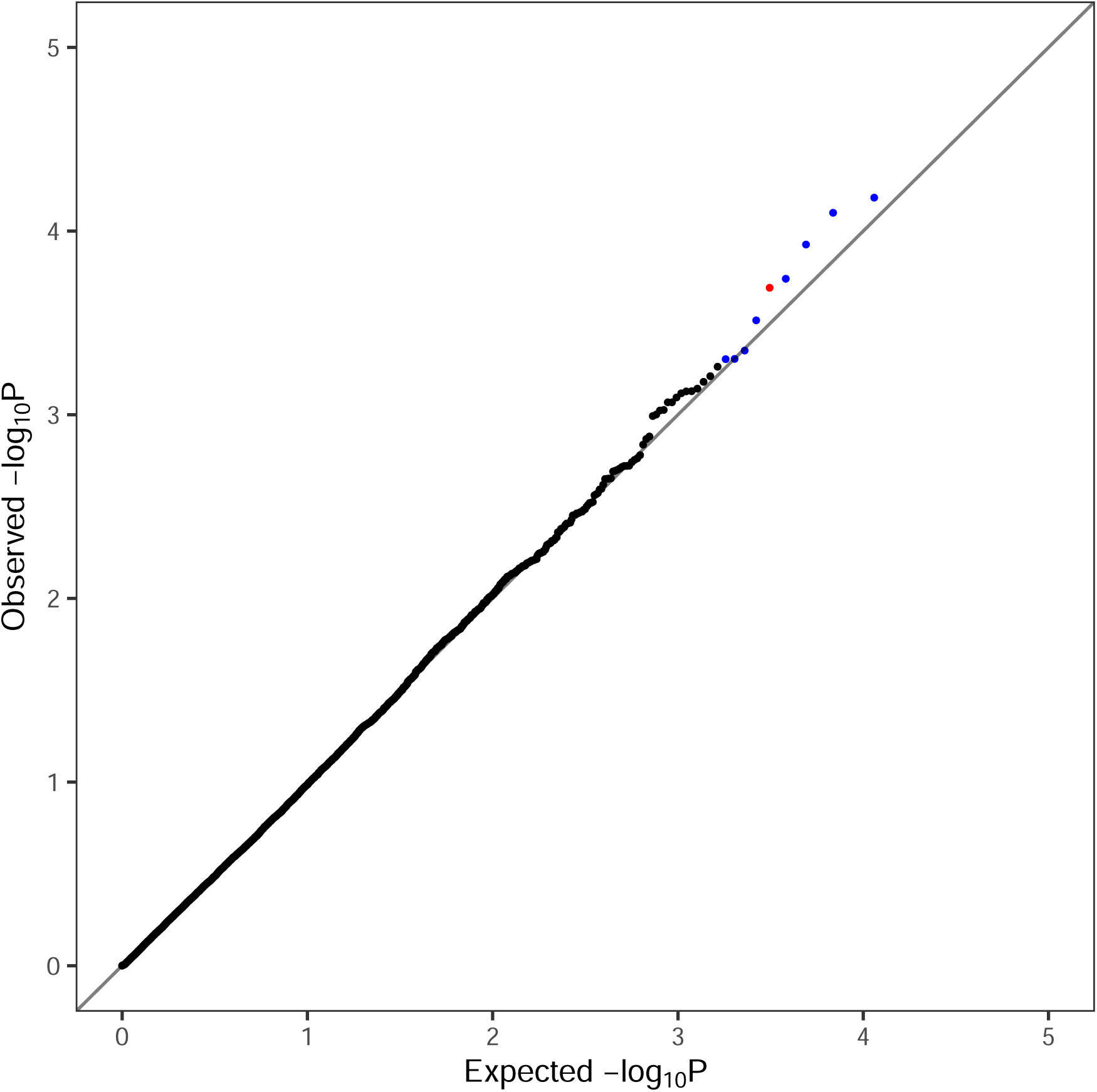
Quantile-Quantile Plot of P-values from assessing the association between rare variant carriers in 5’ UTR regions and breast cancer risk using the burden test. The x-axis is the expected log_10_ P-values from the null hypothesis, the y-axis is the observed log_10_ P-values. Blue dots correspond to regions associated with increased risk and red dots correspond to regions associated with decreased risk. All P-values are unadjusted for multiple testing.

**Figure 3:**
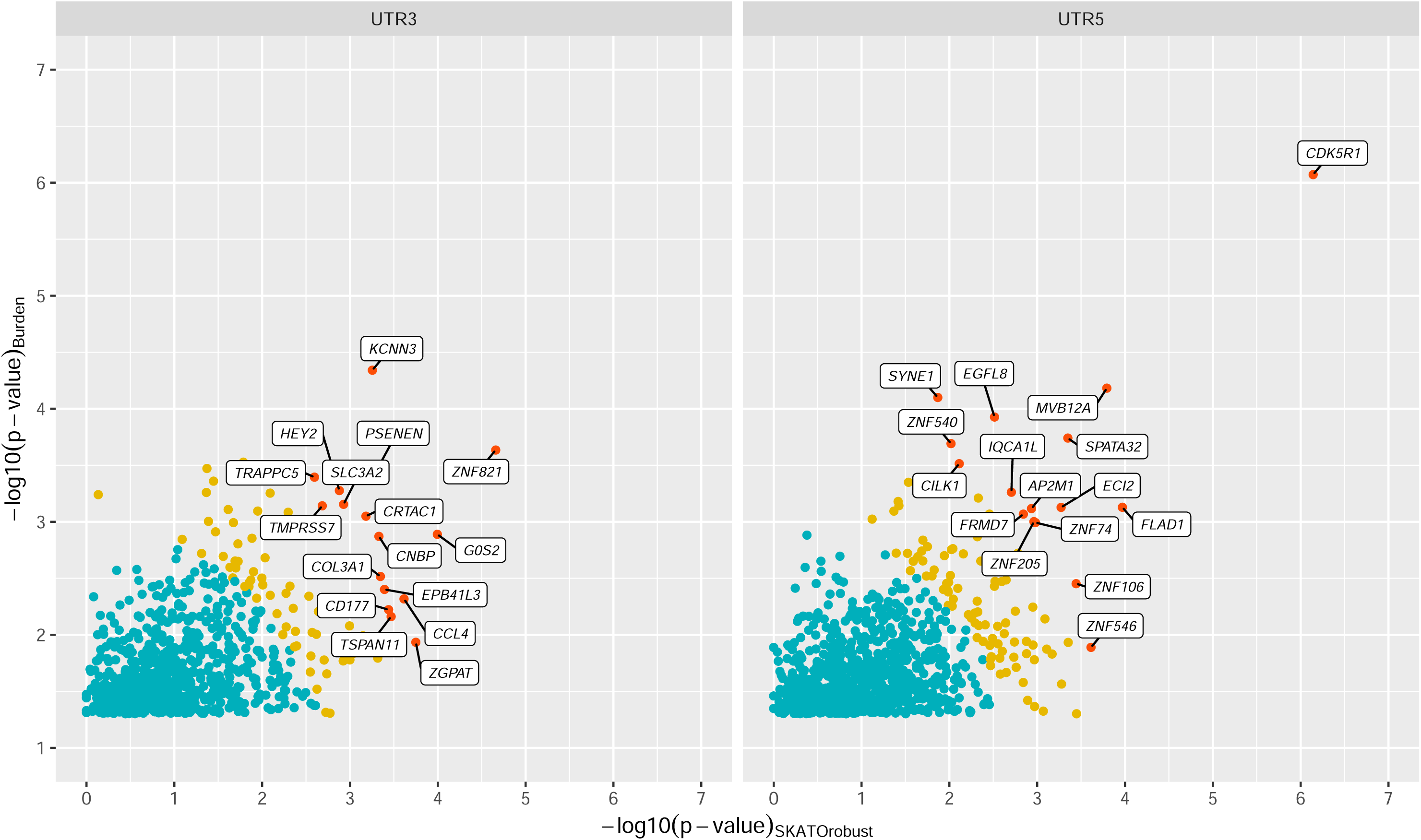
Scatter Plot comparing P-values from the burden test and the robust SKAT-O test for 3’ and 5’ UTR regions for regions with burden test P-value<0.05. The x-axis is the −ve log_10_ P-values from the robust SKAT-O test (P_S_) and the y-axis is the −ve log_10_ P-values from the burden test (P_B_). Genes with P-value<0.001 for either the burden or robust SKAT-O test are labelled. Blue dots correspond to (−*log*_10_ (*P_B_*))^2^ + (−*log*_10_ (*P_S_*))^2^ ≤ 9, orange corresponds to 9< (−*log*_10_ (*P_B_*))^2^ + (−*log*_10_ (*P_S_*))^2^ ≤ 16 and red correspond to (−*log*_10_ (*P_B_*))^2^ + (−*log*_10_ (*P_S_*))^2^ ≥ 16. All P-values are unadjusted for multiple testing.

The most significant association, meeting P<10^−6^ for both the burden and robust-SKAT-O test, was *CDK5R1*. UTR variants were in two distinct regions approximately 350bp apart, with 69 variants with positions 32486993-32487160 and 36 variants with positions 32487483-32487620 (Figure 4). The effect sizes did not differ significantly between the two regions (Supplementary Table 5). The interval between the UTR regions, 32487172-32487467 was an intron, which had no association with risk (Supplementary Table 5). A table of the UTR variants in *CDK5R1*, sorted by the number of carriers, is provided in Supplementary Table 6. There were 9 variants at position 32487007 with varying repeats of GCC (combined P_B_=0.0030, P_S_=0.0017).

**Figure 4:**
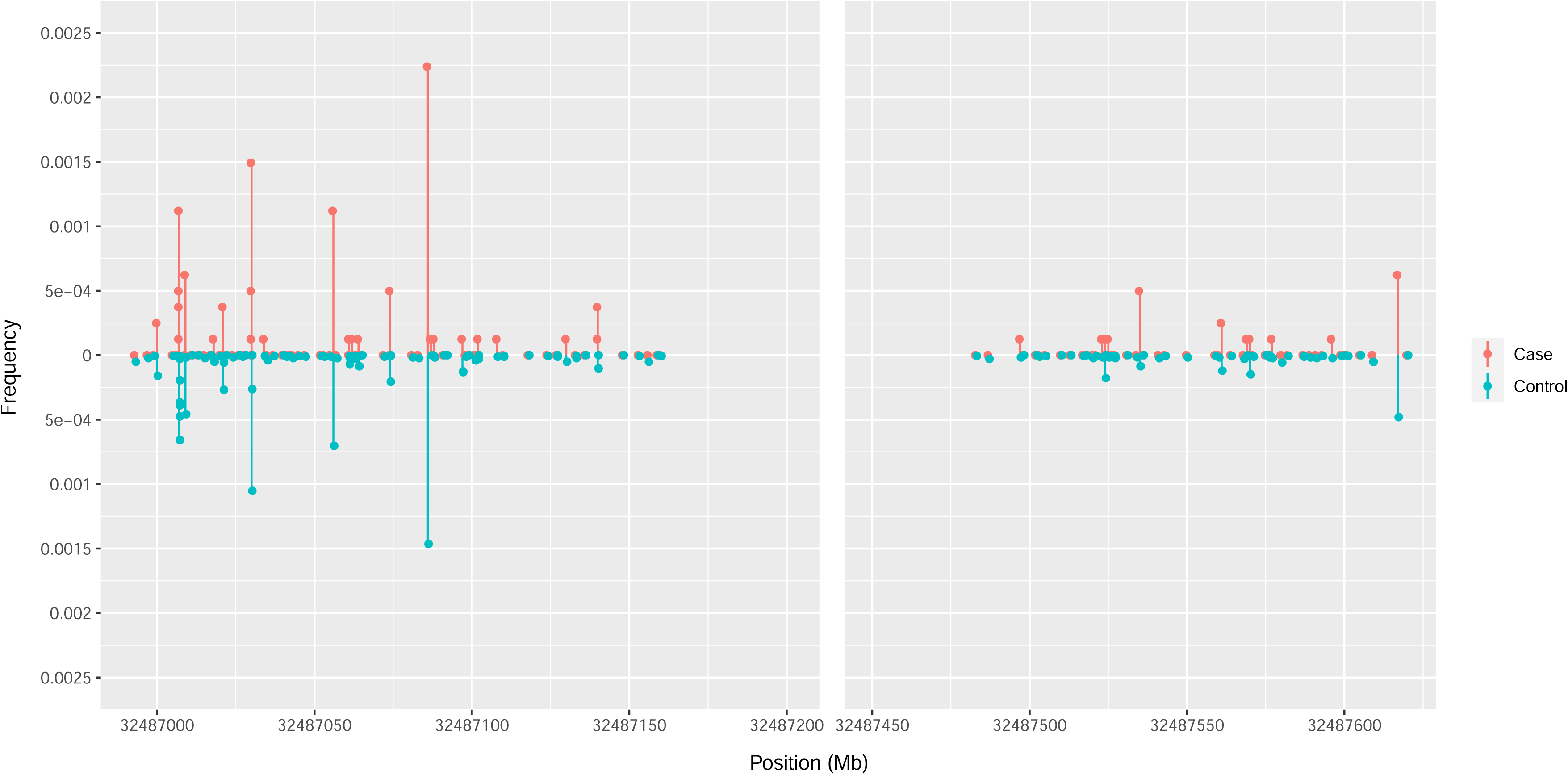
Lollipop Plot of the frequency and position of 5’ UTR variants for *CDK5R1*. The x-axis is the chromosomal position, and the y-axis is the frequency for cases (red; above y=0) and controls (blue; below y=0). The frequency is calculated for females in the dataset only.

### 3’ UTRs

18 3’ UTR regions were associated with breast cancer risk at P_B_<0.001 (Figures 5 and 6; Supplementary Table 7), similar to the number (16) expected by chance. 11 of these corresponded to an increased risk, compared with ~ 8 which would be expected by chance. SKAT-O robust was tested on 821 3’ UTR regions with P_B_<0.05; 17 of these had associations with P_S_<0.001 (Figure 3, Supplementary Table 8). Associations with P_B_ <1×10^−4^ or P_S_ <1×10^−4^ were observed for: *KCNN3* (P_B_ =4.6×10^−5^, P_S_ =5.6×10^−4^) and *ZNF821* (P_B_ =2.3×10^−4^, P_B_ =2.2×10^−5^). None of the five most clearly established breast cancer risk genes (*ATM, BRCA1, BRCA2, CHEK2,* and *PALB2*) had P_B_ or P_S_ <0.001 (Supplementary Table 9).

**Figure 5:**
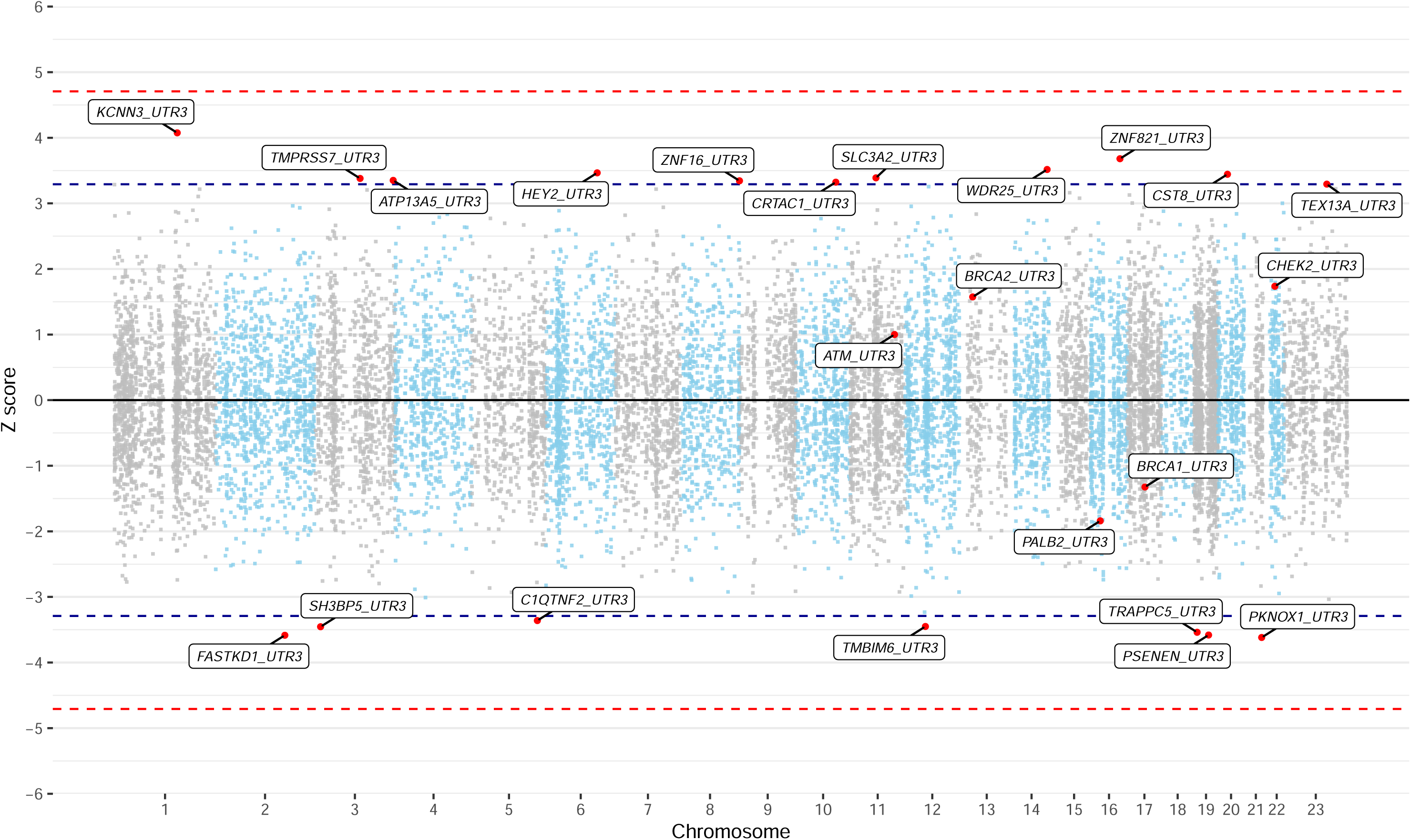
Manhattan Plot of Z-scores from assessing the association between rare variant carriers in 3’ UTR regions and breast cancer risk with the burden test. The x-axis is the chromosomal position, and the y-axis is the Z-score from testing H0 of no association. The blue lines correspond to Z=±3.29, P=0.001, and the red lines correspond to Z=±4.71, P=2.5×10^−6^. All labelled genes are those with P<0.001. P-values are unadjusted for multiple testing.

**Figure 6:**
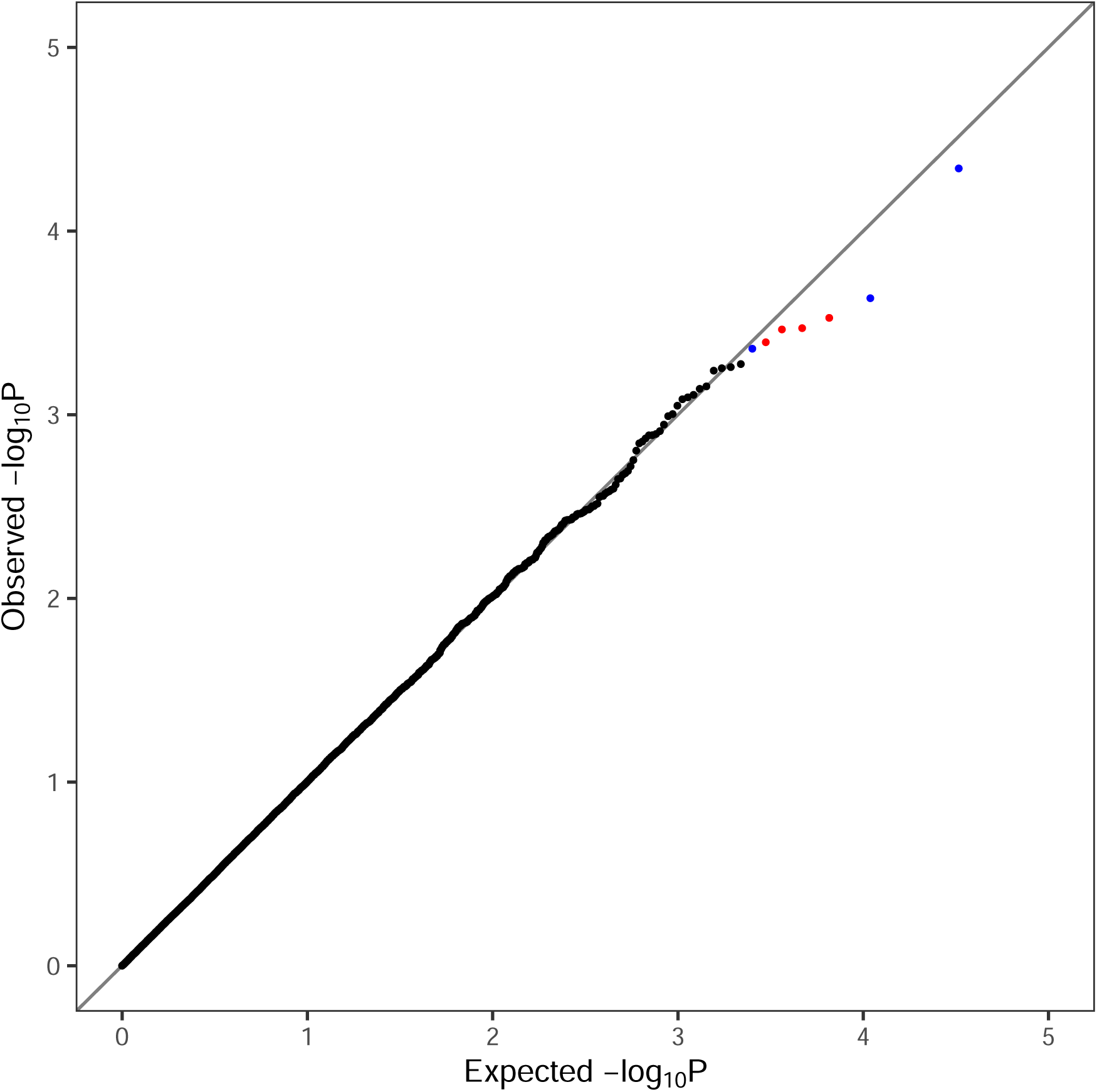
Quantile-Quantile Plot of P-values from assessing the association between rare variant carriers in 3’ UTR regions and breast cancer risk with the burden test. The x-axis is the expected log_10_ P-values from the null hypothesis, the y-axis is the observed log_10_ P-values. Blue dots correspond to regions associated with increased risk and red dots correspond to regions associated with decreased risk. All P-values are unadjusted for multiple testing.

### UTR UK Biobank and BRIDGES meta-analysis

We obtained additional data on UTR regions for 35 putative breast cancer susceptibility genes that were sequenced in the BRIDGES study^1^ and performed a meta-analysis of burden associations across the UK Biobank and BRIDGES datasets. No regions were associated with breast cancer risk at P_BM_<0.001 (Supplementary Table 10). The strongest associations were for the *BARD1* 5’ UTR (P_BM_=0.0076) and *CHEK2* 3’ UTR (P_BM_=0.017).

### Promoters

31 promoter regions were associated at P_B_<0.001 (Figures 7 and 8; Supplementary Table 11), consistent with the number (35.2) expected by chance. However, 23 of the 31 regions at P_B_<0.001 correspond to an increased risk, compared with ~17.6 which would be expected by chance. SKAT-O robust was tested on 1,756 promoter regions with P_B_<0.05; 25 of these had P_S_<0.001 (Figure 9, Supplementary Table 12). For the most significant associations, we identified possible genes associated with each promoter by searching for those with transcription start site (TSS) within 500bp of the promoter end. For each promoter, 1 gene was identified satisfying this condition.

**Figure 7:**
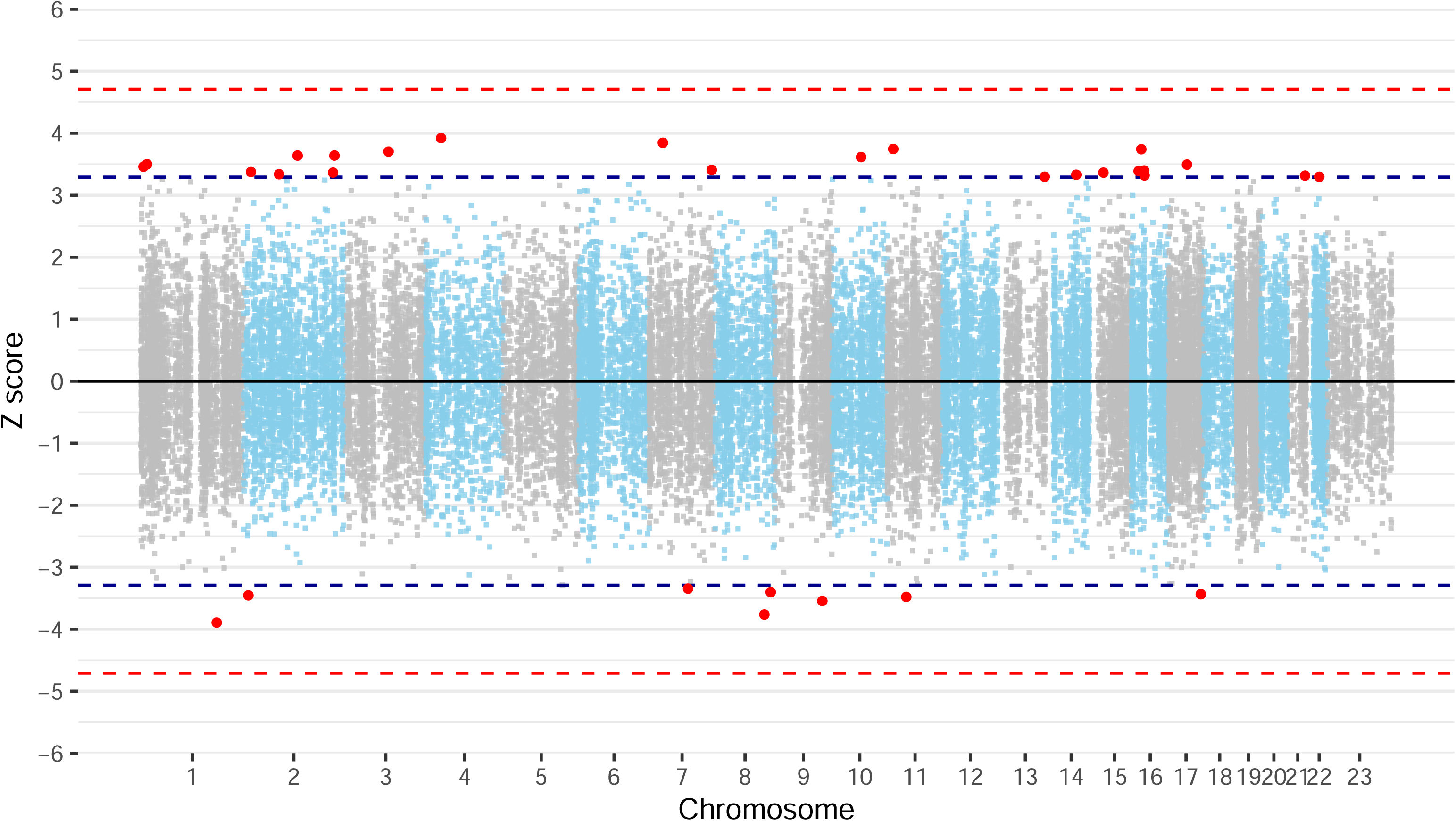
Manhattan Plot of Z-scores from assessing the association between rare variant carriers in promoter regions and breast cancer risk with the burden test. The x-axis is the chromosomal position, and the y-axis is the Z-score from testing H0 of no association. The blue lines correspond to Z=±3.29, P=0.001, and the red lines correspond to Z=±4.71, P=2.5×10^−6^. All red dots are regions with P<0.001. P-values are unadjusted for multiple testing.

**Figure 8:**
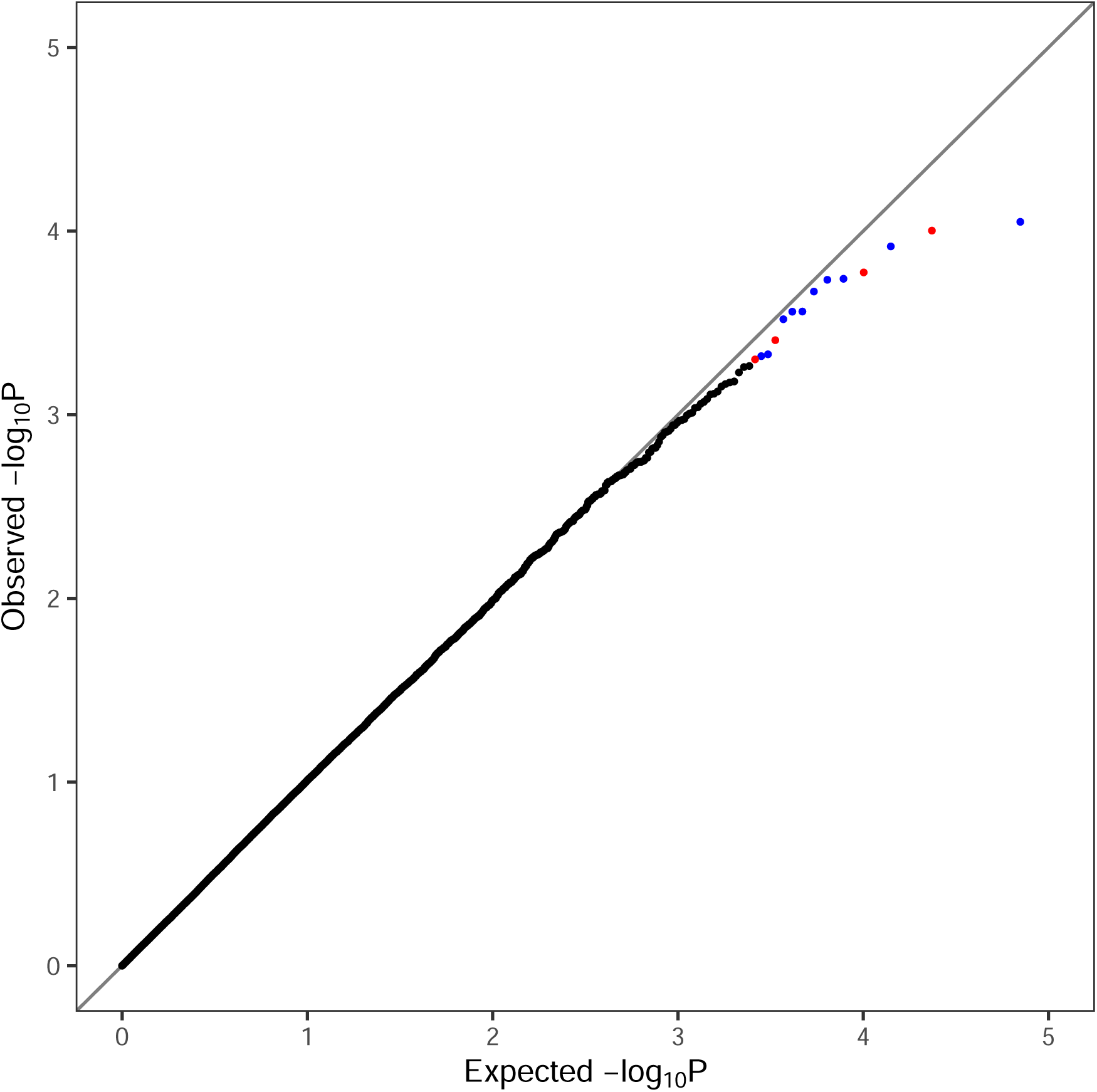
Quantile-Quantile Plot of P-values from assessing the association between rare variant carriers promoter regions and breast cancer risk with the burden test). The x-axis is the expected log_10_ P-values from the null hypothesis, the y-axis is the observed log_10_ P-values. Blue dots correspond to regions associated with increased risk and red dots correspond to regions associated with decreased risk. All P-values are unadjusted for multiple testing.

**Figure 9:**
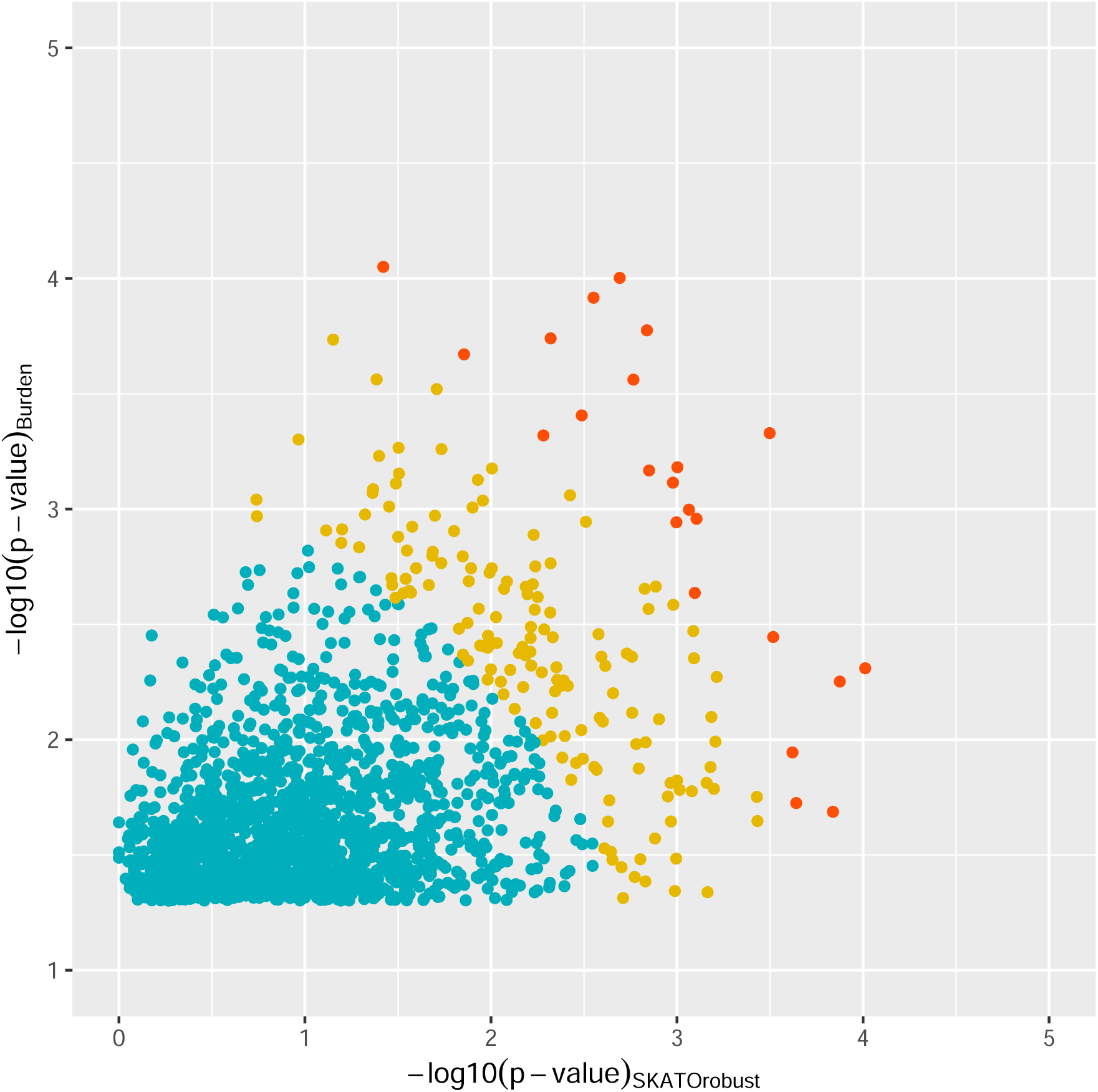
Scatter Plot comparing P-values from burden tests and the robust SKAT-O test for promoter regions for regions with burden test P-value<0.05. The x-axis is the −ve log_10_ P-values from the robust SKAT-O test (P_S_) and the y-axis is the −ve log_10_ P-values from the burden test (P_B_). Genes with P-value<0.001 for either the burden or robust SKAT-O test are labelled. Blue dots correspond to (−*log*_10_ (*P_B_*))^2^ + (−*log*_10_ (*P_S_*))^2^ ≤ 9, orange correspond to 9< (−*log*_10_ (*P_B_*))^2^ + (−*log*_10_ (*P_S_*))^2^ ≤ 16 and red correspond to (−*log*_10_ (*P_B_*))^2^ + (−*log*_10_ (*P_S_*))^2^ ≥ 16. All P-values are unadjusted for multiple testing.

Associations with P_B_ <1×10^−4^ or P_S_ <1×10^−4^ were observed for 3 promoter regions: ENSR00001081062 in 4p14 for with nearest downstream gene *NWD2* (P_B_ =8.9×10^−5^, P_S_ =0.038), ENSR00000016877 in 1q25.3 with nearest downstream gene *ARPC5* (P_B_ =9.9×10^−5^, P_S_ =2.0×10^−3^) and ENSR00000189328 in 5q33.2 with nearest downstream gene *SAP30L* (P_B_ =0.0049, P_S_ =9.7×10^−5^). None of the five most clearly established breast cancer risk genes had P_B_ or P_S_ <0.001 (Supplementary Table 13).

## Discussion

Of the regulatory genomic regions we examined, the strongest association with breast cancer risk was for rare variants in the 5’UTR of *CDK5R1*, which reached P<10^−6^ using both the simple burden and robust SKAT-O tests. *CDK5R1* shows increased expression in many malignancies and has been associated with poor prognosis, proliferation, and drug resistance in many cancers including breast cancer^9^. Of the other 5’UTRs associated at P<0.0001, SYNE1 is a nuclear envelope protein critical for cellular structure and signalling, which is downregulated in many malignancies. Loss of function mutations in the gene have been found in ovarian cancer patients, and downregulation of the gene has been associated with increased tumour mutation burden and immune cell infiltration^10, 11^. Hypermethylation of the *SYNE1* promoter has also been associated with tumour aggressiveness in breast cancer^12^ and poor outcomes in gastric cancer^13^. MVB12A forms part of the ESCRT-1 complex, which has potential links to cancer^14^. Two 3’ UTR associations reached P_B_ <1×10^−4^ or P_S_ <1×10^−4^: *KCNN3* and *ZNF821*. *KCNN3* encodes SK3, also known as KCa2.3, a potassium channel. Increased expression of SK3 is associated with greater breast cancer cell migration^15^. Furthermore, decreased expression levels of *KCNN3* have been associated with drug resistance and poor prognosis for ovarian cancer^16^. ZNF821 interacts with ATM, encoded by the known breast cancer susceptibility gene *ATM* involved in DNA damage signalling and repair^17^.

Three promoter regions reached P_B_ <1×10^−4^ or P_S_ <1×10^−4^: on 1q25.3 (closest likely target *ARPC5*), on 5q23.2 (*SAP30L*) and on 4p14 (*NWD2*), though none reached P<10^−6^. All were high-confidence promoters for these genes, although other genes may be regulated by these promoters. *ARPC5* encodes one of 7 subunits of the Arp2/3 protein complex, which is overexpressed in a variety of cancers including breast cancer^18^. This protein is thought to be involved in the mechanism controlling tumour cell migration, invasion, and metastasis, by mediating actin polymerisation, and therefore to be closely linked to tumour prognosis^18^. CRISPR studies have additionally shown that loss of *ARPC5* can delay the migration of adherent MDA-MB-231 cells (a triple-negative breast cancer cell line)^19^. SAP30L is a SAP30-like protein that, along with SAP30, is a subunit of the SIN3 protein complex, which has suggested roles in breast cancer progression including gene upregulation affecting cell motility, angiogenesis and lymphangiogenesis^20, 21^. *NWD2* is a paralog of *AAMP* which plays a role in angiogenesis and cellular migration. High expression of *AAMP* has been associated with poor prognosis and metastasis of breast cancer^22^.

We considered two types of burden analyses: a simple burden analysis in which all rare variants are considered equivalent, and robust SKAT-O which allows for variation in effect across variants. The latter may be more powerful if only a subset of variants are risk-associated or there are effects in opposite directions, while the simpler test is likely to be more powerful if the effects are similar in magnitude, and also has the advantage that power can be improved by taking family history of the disease into account. Notably, the *CDK5R1* 5’UTR association was the strongest using either method.

Previous GWAS have identified more than 300 common susceptibility loci for breast cancer^23–25^, but few of these have been definitively mapped to UTRs or promoters, with the majority apparently located in more distal regulatory regions. A notable exception is the variant rs78378222 in the 3’UTR of *TP53*, which is associated (in opposite directions) with both ER-negative and ER-positive breast cancer^23^. In this analysis, one association reached P<10^−6^, and the excess of positive associations at P<0.001 suggests that additional breast cancer susceptibility variants may be present in these regions. Many of the putative associations are in regions regulating plausible cancer-related genes, but further, and even larger, replication studies will be required to validate these associations and provide reliable risk estimates.

## Methods

### UK Biobank

UK Biobank is a prospective cohort study of more than 500,000 individuals. Detailed information is given elsewhere^26, 27^. WGS data for 200,000 individuals were released in November 2021 and accessed via the UK Biobank DNA Nexus platform. QC metrics were applied to Variant Call Format files as described by Gardener et al, including genotype level filters for depth and genotype quality^28^. Other filters, including samples with disagreement between genetically determined and self-reported sex and excess relatives, were applied as described elsewhere^2^. Cases were defined by having invasive breast cancer (International Classification of Diseases (ICD)-10 code C50) or carcinoma in situ (D05), as determined by linkage to the National Cancer Registration and Analysis Service (NCRAS), or self-reported breast cancer. Both prevalent and incident cases were included. Only breast cancers that were an individual’s first or second diagnosed cancer were included as cases. By this definition, 8,001 female and 38 male cases were included.

### BRIDGES

For the meta-analysis of UTR regions, we also performed the analysis in the BRIDGES dataset for 35 genes sequenced on a targeted sequencing panel, as described elsewhere^1^. This dataset included 51,494 women with breast cancer and 43,884 women without breast cancer from 43 studies participating in the Breast Cancer Association Consortium (BCAC). Phenotype data were based on the BCAC database v14. Of these 41,609 women with breast cancer were from cohort or population-based case-control studies and unselected for family history. The remainder were from clinic-based studies with some oversampling for familial cases. Details of the sequencing methodology, variant calling and quality control are described in detail elsewhere^1^.

### Data preparation

Promoter regions were extracted using the Ensembl BioMart^5^ data mining tool web page. These regions do not directly identify a specific target gene. To identify the likely corresponding gene for each significant region, we identified genes with TSS within 500bp of the promoter end. UTR regions for genes were similarly extracted from Ensembl BioMart using the R package biomaRt.

Ensemble Variant Effect Predictor (VEP)^6^ v101.0 was used to annotate variants within regions of interest. Annotations included the 1000 genomes phase 3 allele frequency, sequence ontology variant consequences and exon/intron number. For each region, the MANE Select transcript^29^ was used, if available, otherwise the RefSeq Select transcript was used^30^. Annotation files were used to identify rare variants in promoter regions and variants in UTR regions. PTVs, missense and other coding variants were excluded.

### Rare variant analysis

Association analyses were carried out separately for each promoter or UTR region. The main association analyses were burden tests in which genotypes were collapsed into a 0/1 variable based on whether samples carried a variant in the region. For each region we estimated odds ratios and confidence intervals, and derived Z-scores, and two-sided P-values (P_B_), using logistic regression. The method used here also incorporates the family history of breast cancer as a surrogate for disease status with weight ½, as explained in detail elsewhere^2^. This model assumes that all variants are associated with the same effect size and is expected to be most powerful under this assumption. SKAT is a method that allows for different variants within a region to have different effects or no effect and with different effect directions^31^. SKAT is likely to be more powerful when a smaller proportion of variants are causal, or effects have different directions^32^. SKAT-O is an optimal test that is a linear combination of the burden and SKAT statistic, which optimises the weighting of the two tests for each region^26^. In large biobanks with unbalanced case-control ratios, these methods can suffer from inflated type-1 error rates. A robust SKAT-O method was recently developed that accounts for this using Saddle Point Approximation and Efficient Resampling^33^. This method is more computationally intensive than the simple burden test and we therefore only obtain SKAT-O robust P-values (P_S_) for promoter or UTR regions with P_B_<0.05. Exome-wide significance is defined as P<2.5×10^−6^. Here we define a more stringent level at P<10^−6^ given we consider approximately 35,000 promoter regions and 34,000 UTR regions. However, we additionally consider regions with P<0.001 to be of interest.

For promoter regions, 3’ UTR regions and 5’ UTR regions, separate Manhattan plots and QQ plots were made for the Z-scores and P_B_ values from the simple burden tests. For regions with P_B_<0.05 scatter plots summarising P_S_ and P_B_ values were made.

For genes in both the UKB and BRIDGES datasets, we combined Z-scores for the UTR regions in a meta-analysis using an inverse variance weighting approach. The combined Z-score was defined as 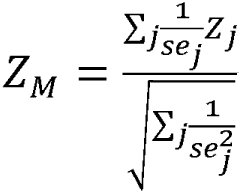. Here, *Z_M_* is the combined z score, *Z_j_* the z-score for study j and *se_j_*, is the standard error of *β_j_* = *log*(*OR*j).

## Statistics and reproducibility

No statistical method was used to predetermine the sample size. The experiments were not randomized, and we did not use blinding. Some samples were excluded for reasons as described in the methods above, for example, for sex discrepancies, excess relatives, or discrepancies with previous genotyping.

## Data Availability

Individual level data for the BRIDGES data are not publicly available due to ethical review board constraints but are available on request through the BCAC Data Access Co-ordinating Committee. Requests for access to UK Biobank data should be made to the UK Biobank Access Management Team.

## Code Availability

Quality Control filtering of vcf files was performed using vcftools v0.1.15, bcftools v1.9, picard v2.22.2 and plink v1.90b, as outlined in the methods. Variants were annotated using Ensembl Variant Effect Predictor v101 with assembly GRCh38. The code for each software is available at the website of each package. Data manipulation and analysis were performed using R-4.3.3 with packages clusterProfiler (4.2.2), data.table (1.14.2), dplyr (1.0.9), dbplyr (2.5.0), gtools (3.9.5), HGNChelper (0.8.9), SKAT (2.2.5), tibble (3.2.1) and tidyr (1.3.1). Plots were created using additional packages ggplot2 (3.5.1) and ggrepel (0.9.5). The code for each of the R packages can be found in their associated vignettes.

## Data Availability

Data from UK Biobank are available through application to UK Biobank. Data from the Breast Cancer Association Consortium (BCAC) used in the present study are available upon reasonable request through the BCAC Data Access Coordinating Committee. All data produced in the present study are available upon reasonable request to the authors

## Acknowledgments

The research has been conducted using the UK Biobank Resource under Application Number 28126. N.W. was supported by the International Alliance for Cancer Early Detection, an alliance between Cancer Research UK (C14478/A29329), Canary Center at Stanford University, the University of Cambridge, OHSU Knight Cancer Institute, University College London, and the University of Manchester. J.P.T. was supported by Cancer Research UK (PRCPJT-May21\100006 and G110748). Quality control of the UK Biobank sequencing data has been funded by the Medical Research Council (unit programs: MC_UU_12015/2, MC_UU_00006/2). The BRIDGES project was supported by the European Union Horizon 2020 research and innovation programs BRIDGES (grant number, 634935 to P.D A.G.-N., A,M.D. and D.F.E) and B-CAST (633784 to M.K.S. and D.F.E.), the Wellcome Trust (v203477/Z/16/Z to S.H.T ad D.F.E), and Cancer Research UK (C1287/A16563). Details regarding funding of specific BRIDGES studies are provided in the Supplementary Material.

## Author contributions

D.F.E. supervised this work and directed the overall analysis. N.W. performed the statistical analysis. N.W., J.P.T., L.D., J.D., M.N, E.J.G. and J.R.B.P., developed the bioinformatics and computational pipelines. M.K.B., S.B., R.K. and Q.W. led data management within the BCAC. J.C-C. and M.K.S. led working groups within the BCAC. A.M.D. and P.D. directed the BRIDGES project. M.A., T.U.A., I.L. A., A.C.A., S.E.B., M.K.B., H.B., N.J.C., J.C.-C., K.C., T.D., D.G.E., P.A.F., J.D.F., H.F., A.G.-N., P.G., E.H., P. H., M.H,, M.J.H., A.J., V.N.K., J.L., A.Lindblom, A.Lophatananon, A.M., S.M., R.L. M., N.O., M.I.P., S.K.P., M.U.R., E.S., E.J.S., M.K.S., M.C.S., A.B.S., D.T., Q.W., J.S. and S.H.T. contributed to the design and conduct of the contributing BCAC studies.

N.W. and D.F.E. drafted the manuscript. All authors reviewed and approved the paper.

