## Supplementary Material for "Analysis of variants in untranslated and promoter regions and breast cancer risk using whole genome sequencing data"

### Supplementary Note

#### BCAC study-specific funding

This work was supported by Cancer Research UK grant: PPRPGM-Nov20\100002 and by core funding from the NIHR Cambridge Biomedical Research Centre (NIHR203312) [*]. *The views expressed are those of the author(s) and not necessarily those of the NIHR or the Department of Health and Social Care. Additional funding for BCAC is provided by the Confluence project which is funded with intramural funds from the National Cancer Institute Intramural Research Program, National Institutes of Health, the European Union's Horizon 2020 Research and Innovation Programme (grant numbers 634935 and 633784 for BRIDGES and B-CAST respectively), and the PERSPECTIVE I&I project, funded by the Government of Canada through Genome Canada and the Canadian Institutes of Health Research, the Ministère de l'Économie et de l'Innovation du Québec through Genome Québec, the Quebec Breast Cancer Foundation. The EU Horizon 2020 Research and Innovation Programme funding source had no role in study design, data collection, data analysis, data interpretation or writing of the report.

The BRIDGES panel sequencing was supported by the European Union Horizon 2020 research and innovation program BRIDGES (grant number 634935) and the Wellcome Trust (v203477/Z/16/Z).

The Australian Breast Cancer Family Study (ABCFS) was supported by grant UM1 CA164920 from the National Cancer Institute (USA). The content of this manuscript does not necessarily reflect the views or policies of the National Cancer Institute or any of the collaborating centres in the Breast Cancer Family Registry (BCFR), nor does mention of trade names, commercial products, or organizations imply endorsement by the USA Government or the BCFR. The ABCFS was also supported by the National Health and Medical Research Council of Australia, the New South Wales Cancer Council, the Victorian Health Promotion Foundation (Australia) and the Victorian Breast Cancer Research Consortium. J.L.H. is a National Health and Medical Research Council (NHMRC) Senior Principal Research Fellow. M.C.S. is a NHMRC Senior Research Fellow. The ABCS study was supported by the Dutch Cancer Society [grants NKI 2007-3839; 2009 4363] and an institutional grant of the Dutch Cancer Society and the Dutch Ministry of Health, Welfare and Sport. The Australian Breast Cancer Tissue Bank (ABCTB) was supported by the National Health and Medical Research Council of Australia, The Cancer Institute NSW and the National Breast Cancer Foundation. The ACP study is funded by the Breast Cancer Research Trust, UK. KM and AL are supported by the NIHR Manchester Biomedical Research Centre, the Allan Turing Institute under the EPSRC grant EP/N510129/1. The work of the BBCC was partly funded by ELAN-Fond of the University Hospital of Erlangen. The BBCC is funded by Cancer Research UK and Breast Cancer Now and acknowledges NHS funding to the NIHR Biomedical Research Centre, and the National Cancer Research Network (NCRN). For BIGGS, ES is supported by NIHR Comprehensive Biomedical Research Centre, Guy's & St. Thomas' NHS Foundation Trust in partnership with King's College London, United Kingdom. It is supported by the Oxford Biomedical Research Centre. BOCS is supported by funds from Cancer Research UK (C8620/A8372/A15106) and the Institute of Cancer Research (UK). BOCS acknowledges NHS funding to the Royal Marsden / Institute of Cancer Research NIHR Specialist Cancer Biomedical Research Centre. The BREast Oncology GAlician Network (BREOGAN) is funded by Acción Estratégica de Salud del Instituto de Salud Carlos III FIS PI12/02125/Cofinanciado and FEDER PI17/00918/Cofinanciado FEDER; Acción Estratégica de Salud del Instituto de

Salud Carlos III FIS Intrasalud (PI13/01136); Programa Grupos Emergentes, Cancer Genetics Unit, Instituto de Investigacion Biomedica Galicia Sur. Xerencia de Xestion Integrada de Vigo-SERGAS, Instituto de Salud Carlos III, Spain; Grant 10CSA012E, Consellería de Industria Programa Sectorial de Investigación Aplicada, PEME I + D e I + D Suma del Plan Gallego de Investigación, Desarrollo e Innovación Tecnológica de la Consellería de Industria de la Xunta de Galicia, Spain; Grant EC11-192. Fomento de la Investigación Clínica Independiente, Ministerio de Sanidad, Servicios Sociales e Igualdad, Spain; and Grant FEDER-Innterconecta. Ministerio de Economia y Competitividad, Xunta de Galicia, Spain. The BSUCH study was supported by the Dietmar-Hopp Foundation, the Helmholtz Society and the German Cancer Research Center (DKFZ). The CAMA study was funded by Consejo Nacional de Ciencia y Tecnología (CONACyT) (SALUD-2002-C01-7462). Sample collection and processing was funded in part by grants from the National Cancer Institute (NCI R01CA120120 and K24CA169004). CCGP is supported by funding from the University of Crete. The CECILE study was supported by Fondation de France, Institut National du Cancer (INCa), Ligue Nationale contre le Cancer, Agence Nationale de Sécurité Sanitaire, de l'Alimentation, de l'Environnement et du Travail (ANSES), Agence Nationale de la Recherche (ANR). The CGPS was supported by the Chief Physician Johan Boserup and Lise Boserup Fund, the Danish Medical Research Council, and Herlev and Gentofte Hospital. The CNIO-BCS was supported by the Instituto de Salud Carlos III, the Red Temática de Investigación Cooperativa en Cáncer and grants from the Asociación Española Contra el Cáncer and the Fondo de Investigación Sanitario (PI11/00923 and PI12/00070). COLBCCC is supported by the German Cancer Research Center (DKFZ), Heidelberg, Germany. Diana Torres was in part supported by a postdoctoral fellowship from the Alexander von Humboldt Foundation. FHRISK and PROCAS are funded from NIHR grant PGfAR 0707-10031. DGE, AH and WGN are supported by the NIHR Manchester Biomedical Research Centre (IS-BRC-1215-20007). The GC-HBOC (German Consortium of Hereditary Breast and Ovarian Cancer) is supported by the German Cancer Aid (grant no 110837 and 70114178, coordinator: Rita K. Schmutzler, Cologne) and the Federal Ministry of Education and Research, Germany (grant no 01GY1901). This work was also funded by the European Regional Development Fund and Free State of Saxony, Germany (LIFE - Leipzig Research Centre for Civilization Diseases, project numbers 713-241202, 713-241202, 14505/2470, 14575/2470). The GENICA was funded by the Federal Ministry of Education and Research (BMBF) Germany grants 01KW9975/5, 01KW9976/8, 01KW9977/0 and 01KW0114, the Robert Bosch Foundation, Stuttgart, Deutsches Krebsforschungszentrum (DKFZ), Heidelberg, the Institute for Prevention and Occupational Medicine of the German Social Accident Insurance, Institute of the Ruhr University Bochum (IPA), Bochum, as well as the Department of Internal Medicine, Johanniter GmbH Bonn, Johanniter Krankenhaus, Bonn, Germany. Generation Scotland (GENSCOT) received core support from the Chief Scientist Office of the Scottish Government Health Directorates [CZD/16/6] and the Scottish Funding Council [HR03006]. Genotyping of the GS: SFHS samples was carried out by the Genetics Core Laboratory at the Edinburgh Clinical Research Facility, University of Edinburgh, Scotland and was funded by the Medical Research Council UK and the Wellcome Trust (Wellcome Trust Strategic Award “STratifying Resilience and Depression Longitudinally” (STRADL) Reference 104036/Z/14/Z). Funding for identification of cases and contribution to BCAC funded in part by the Wellcome Trust Seed Award “Temporal trends in incidence and mortality of molecular subtypes of breast cancer to inform public health, policy and prevention” Reference 207800/Z/17/Z. The GEPARSIXTO study was conducted by the German Breast Group GmbH. The GESBC was supported by the Deutsche

Krebshilfe e. V. [70492] and the German Cancer Research Center (DKFZ). GLACIER was supported by Breast Cancer Now, CRUK and the Biomedical Research Centre at Guy's and St Thomas' NHS Foundation Trust and King's College London. The HABCS study was supported by the German Research Foundation (DFG Do761/15-1), the Claudia von Schilling Foundation for Breast Cancer Research, by the Lower Saxonian Cancer Society, and by the Rudolf Bartling Foundation The HEBON study is supported by the Dutch Cancer Society grants NKI1998-1854, NKI2004-3088, NKI2007-3756, the Netherlands Organisation of Scientific Research grant NWO 91109024, the Pink Ribbon grants 110005 and 2014-187.WO76, the BBMRI grant NWO 184.021.007/CP46 and the Transcan grant JTC 2012 Cancer 12-054. The HMBCS was supported by the German Research Foundation (DFG Do761/15-1), a grant from the Friends of Hannover Medical School, and by the Rudolf Bartling Foundation. The HUBCS was supported by the German Research Foundation (DFG Do761/15-1), a grant from the German Federal Ministry of Research and Education (RUS08/017), B.M. was supported by grant 17-44-020498, 17-29-06014 of the Russian Foundation for Basic Research, D.P. was supported by grant 18-29-09129 of the Russian Foundation for Basic Research, E.K was supported by the mega grant from the Government of Russian Federation (2020-220-08-2197), and the study was performed as part of the assignment of the Ministry of Science and Higher Education of the Russian Federation (№AAAA-A16-116020350032-1). Financial support for KARBAC was provided through the regional agreement on medical training and clinical research (ALF) between Stockholm County Council and Karolinska Institutet, the Swedish Cancer Society, The Gustav V Jubilee Foundation and Bert von Kantzows Foundation. The KARMA study was supported by Märit and Hans Rausings Initiative Against Breast Cancer. The KBCP was financially supported by the special Government Funding (VTR) of Kuopio University Hospital grants, the Cancer Fund of North Savo, the Finnish Cancer Organizations, and the strategic funding of the University of Eastern Finland. kConFab is supported by a grant from the National Breast Cancer Foundation, and previously by the National Health and Medical Research Council (NHMRC), the Queensland Cancer Fund, the Cancer Councils of New South Wales, Victoria, Tasmania and South Australia, and the Cancer Foundation of Western Australia. Financial support for the AOCS was provided by the United States Army Medical Research and Materiel Command [DAMD17-01-1-0729], Cancer Council Victoria, Queensland Cancer Fund, Cancer Council New South Wales, Cancer Council South Australia, The Cancer Foundation of Western Australia, Cancer Council Tasmania and the National Health and Medical Research Council of Australia (NHMRC; 400413, 400281, 199600). G.C.T. and P.W. are supported by the NHMRC. RB was a Cancer Institute NSW Clinical Research Fellow. The KOHBRA study was partially supported by a grant from the Korea Health Technology R&D Project through the Korea Health Industry Development Institute (KHIDI), and the National R&D Program for Cancer Control, Ministry of Health & Welfare, Republic of Korea (HI16C1127; 0720450; 1020350; 1420190). The MARIE study was supported by the Deutsche Krebshilfe e.V. [70-2892-BR I, 106332, 108253, 108419, 110826, 110828], the Hamburg Cancer Society, the German Cancer Research Center (DKFZ) and the Federal Ministry of Education and Research (BMBF) Germany [01KH0402]. The MASTOS study was supported by "Cyprus Research Promotion Foundation" grants 0104/13 and 0104/17, and the Cyprus Institute of Neurology and Genetics. MBCSG is supported by grants from the Italian Association for Cancer Research (AIRC). The MCBCSG was supported by the NIH grants R35CA253187, R01CA192393, R01CA116167, R01CA176785 a NIH Specialized Program of Research Excellence (SPORE) in Breast Cancer [P50CA116201], and the Breast Cancer Research

Foundation. The Melbourne Collaborative Cohort Study (MCCS) cohort recruitment was funded by VicHealth and Cancer Council Victoria. The MCCS was further augmented by Australian National Health and Medical Research Council grants 209057, 396414 and 1074383 and by infrastructure provided by Cancer Council Victoria. Cases and their vital status were ascertained through the Victorian Cancer Registry and the Australian Institute of Health and Welfare, including the Australian Cancer Database. MYBRCA is funded by research grants from the Wellcome Trust (v203477/Z/16/Z), the Malaysian Ministry of Higher Education (UM.C/HlR/MOHE/06) and Cancer Research Malaysia. The NBCS has received funding from the K.G. Jebsen Centre for Breast Cancer Research; the Research Council of Norway grant 193387/V50 (to A-L Børresen-Dale and V.N. Kristensen) and grant 193387/H10 (to A-L Børresen-Dale and V.N. Kristensen), South Eastern Norway Health Authority (grant 39346 to A-L Børresen-Dale) and the Norwegian Cancer Society (to A-L Børresen-Dale and V.N. Kristensen). The Ontario Familial Breast Cancer Registry (OFBCR) was supported by grant U01CA164920 from the USA National Cancer Institute of the National Institutes of Health. The content of this manuscript does not necessarily reflect the views or policies of the National Cancer Institute or any of the collaborating centres in the Breast Cancer Family Registry (BCFR), nor does mention of trade names, commercial products, or organizations imply endorsement by the USA Government or the BCFR. The ORIGO study was supported by the Dutch Cancer Society (RUL 1997-1505) and the Biobanking and Biomolecular Resources Research Infrastructure (BBMRI-NL CP16). The PBCS was funded by Intramural Research Funds of the National Cancer Institute, Department of Health and Human Services, USA. Genotyping for PLCO was supported by the Intramural Research Program of the National Institutes of Health, NCI, Division of Cancer Epidemiology and Genetics. The PLCO is supported by the Intramural Research Program of the Division of Cancer Epidemiology and Genetics and supported by contracts from the Division of Cancer Prevention, National Cancer Institute, National Institutes of Health. The RBCS was funded by the Dutch Cancer Society (DDHK 2004-3124, DDHK 2009-4318). The SASBAC study was supported by funding from the Agency for Science, Technology and Research of Singapore (A*STAR), the US National Institute of Health (NIH) and the Susan G. Komen Breast Cancer Foundation. SEARCH is funded by Cancer Research UK [C490/A10124, C490/A16561] and supported by the UK National Institute for Health Research Biomedical Research Centre at the University of Cambridge. The University of Cambridge has received salary support for PDPP from the NHS in the East of England through the Clinical Academic Reserve. SGBCC is funded by the National Research Foundation Singapore, NUS start-up Grant, National University Cancer Institute Singapore (NCIS) Centre Grant, Breast Cancer Prevention Programme, the Asian Breast Cancer Research Fund and the NMRC Clinician Scientist Award (SI Category). Population-based controls were from the Multi-Ethnic Cohort (MEC) funded by grants from the Ministry of Health, Singapore, National University of Singapore and National University Health System, Singapore. SKKDKFZS is supported by the DKFZ. The SMC is funded by the Swedish Cancer Foundation and the Swedish Research Council (VR 2017-00644) grant for the Swedish Infrastructure for Medical Population-based Life-course Environmental Research (SIMPLER). The SZBCS was supported by Grant PBZ_KBN_122/P05/2004 and the program of the Minister of Science and Higher Education under the name "Regional Initiative of Excellence" in 2019-2022 project number 002/RID/2018/19 amount of financing 12 000 000 PLN. Data collection for UBCS was supported by the Utah Population Database (UPDB) and Utah Cancer Registry (UCR). The UPDB is supported by HCI, the University of Utah, and NCI grant P30 CA2014. The UCR is additionally funded by the NCI's SEER Program,

HHSN261201800016I, and the US Center for Disease Control and Prevention's National Program of Cancer Registries (NU58DP007131). The UCIBCS component of this research was supported by the NIH [CA58860, CA92044] and the Lon V Smith Foundation [LVS39420]. The UKBGS is funded by Breast Cancer Now and the Institute of Cancer Research (ICR), London. ICR acknowledges NHS funding to the NIHR Biomedical Research Centre. A.B.S. was supported by an NHMRC Investigator Fellowship (APP177524).

#### BCAC Study-specific acknowledgements

We thank all the individuals who took part in these studies and all the researchers, clinicians, technicians and administrative staff who have enabled this work to be carried out. ABCFS thanks Maggie Angelakos, Judi Maskiell, and Gillian Dite. ABCS thanks the Blood bank Sanquin, The Netherlands. The ACP study wishes to thank the participants in the Thai Breast Cancer study. Special thanks also go to the Thai Ministry of Public Health (MOPH), doctors and nurses who helped with the data collection process. Finally, the study would like to thank Dr Prat Boonyawongviroj, the former Permanent Secretary of MOPH and Dr Pornthep Siriwanarungsan, the former Department Director-General of Disease Control who have supported the study throughout. BBCS thanks Eileen Williams, Elaine Ryder-Mills, and Kara Sargus. BCEES thanks Allyson Thomson, Christobel Saunders, Jennifer Girschik, Jane Heyworth and Terry Boyle. BIGGS thanks Niall McInerney, Gabrielle Colleran, Andrew Rowan, and Angela Jones. The BREOGAN study would not have been possible without the contributions of the following: Manuela Gago-Dominguez, Jose Esteban Castelao, Angel Carracedo, Victor Muñoz Garzón, Alejandro Novo Domínguez, Maria Elena Martinez, Sara Miranda Ponte, Carmen Redondo Marey, Maite Peña Fernández, Manuel Enguix Castelo, Maria Torres, Manuel Calaza (BREOGAN), José Antúnez, Máximo Fraga and the staff of the Department of Pathology and Biobank of the University Hospital Complex of Santiago-CHUS, Instituto de Investigación Sanitaria de Santiago, IDIS, Xerencia de Xestion Integrada de Santiago-SERGAS; Joaquín González-Carreró and the staff of the Department of Pathology and Biobank of University Hospital Complex of Vigo, Instituto de Investigacion Biomedica Galicia Sur, SERGAS, Vigo, Spain. The BSUCH study acknowledges the Principal Investigator, Barbara Burwinkel, and, thanks Peter Bugert, Medical Faculty Mannheim. CCGP thanks Styliani Apostolaki, Anna Margiolaki, Georgios Nintos, Maria Perraki, Georgia Saloustrou, Georgia Sevastaki, Konstantinos Pompodakis. CGPS thanks staff and participants of the Copenhagen General Population Study. For the excellent technical assistance: Dorthe Uldall Andersen, Maria Birna Arnadottir, Anne Bank, Dorthe Kjeldgård Hansen. The Danish Cancer Biobank is acknowledged for providing infrastructure for the collection of blood samples for the cases. The Danish Breast Cancer Cooperative Group (DBCG) are acknowledged for their provision of clinical case data. CNIO-BCS thanks Guillermo Pita, Charo Alonso, Nuria Álvarez, Pilar Zamora, Primitiva Menendez, the Human Genotyping-CEGEN Unit (CNIO). COLBCCC thanks all patients, the physicians Justo G. Olaya, Mauricio Tawil, Lilian Torregrosa, Elias Quintero, Sebastian Quintero, Claudia Ramírez, José J. Caicedo, and Jose F. Robledo, and the technician Michael Gilbert for their contributions and commitment to this study. FHRISK and PROCAS thank NIHR for funding. The GENICA Network: Dr. Margarete Fischer-Bosch-Institute of Clinical Pharmacology, Stuttgart, and the University of Tübingen, Germany [HB, Reiner Hoppe, Wing-Yee Lo], Department of Internal Medicine, Johanniter GmbH Bonn, Johanniter Krankenhaus, Bonn, Germany [Yon Dschun Ko, Christian Baisch], Institute of Pathology, University of Bonn, Germany [Hans-Peter Fischer], Molecular Genetics of Breast Cancer,

Deutsches Krebsforschungszentrum (DKFZ), Heidelberg, Germany [Ute Hamann], Institute for Prevention and Occupational Medicine of the German Social Accident Insurance, Institute of the Ruhr University Bochum (IPA), Bochum, Germany [Thomas Brüning, Beate Pesch, Sylvia Rabstein, Anne Lotz]; and Institute of Occupational Medicine and Maritime Medicine, University Medical Center Hamburg-Eppendorf, Germany [Volker Harth]. HABCS thanks Peter Schürmann, Peter Hillemanns, Natalia Bogdanova, Michael Bremer, Johann Karstens, Hans Christiansen and the Breast Cancer Network in Lower Saxony for continuous support. HEBON Investigators are J. Margriet Collée, Frans B. L. Hogervorst, Maartje J. Hooning, Carolien M. Kets, Peter Devilee, Christi J. van Asperen, Matti A. Rookus, Marjanka K. Schmidt, Cora M. Aalfs, Muriel A. Adank, Margreet G. E. M. Ausems, Marinus J. Blok, Encarna B. Gómez Garcia, Bernadette A. M. Heemskerk-Gerritsen, Antoinette Hollestelle, Agnes Jager, Linetta B. Koppert, Marco Koudijs, Mieke Kriege, Hanne E. J. Meijers-Heijboer, Arjen R. Mensenkamp, Thea M. Mooij, Jan C. Oosterwijk, Ans M. W. van den Ouweland, Frederieke H. van der Baan, Annemieke H. van der Hout, Lizet E. van der Kolk, Rob B. van der Luijt, Carolien H. M. van Deurzen, Helena C. van Doorn, Klaartje van Engelen, Liselotte P. van Hest, Theo A. M. van Os, Senno Verhoef, Maartje J. Vogel & Juul T. Wijnen. HMBCS thanks Peter Hillemanns, Hans Christiansen and Johann H. Karstens. HUBCS thanks Darya Prokofyeva and Shamil Gantsev. KARMA and SASBAC thank the Swedish Medical Research Counsel. KBCP thanks Eija Myöhänen. kConFab/AOCS wish to thank Heather Thorne, Eveline Niedermayr, all the kConFab research nurses and staff, the heads and staff of the Family Cancer Clinics, and the Clinical Follow-Up Study (which has received funding from the NHMRC, the National Breast Cancer Foundation, Cancer Australia, and the National Institute of Health (USA)) for their contributions to this resource, and the many families who contribute to kConFab. We thank all investigators of the KOHBRA (Korean Hereditary Breast Cancer) Study. MARIE thanks Petra Seibold, Nadia Obi, Sabine Behrens, Ursula Eilber and Muhabbet Celik. MASTOS thanks all the study participants and expresses appreciation to the doctors: Yiola Marcou, Eleni Kakouri, Panayiotis Papadopoulos, Simon Malas and Maria Daniel, as well as to all the nurses and volunteers who provided valuable help towards the recruitment of the study participants. MBCSG (Milan Breast Cancer Study Group): Paolo Radice, Paolo Peterlongo, Siranoush Manoukian, Bernard Peissel, Jacopo Azzollini, Erica Rosina, Daniela Zaffaroni, Bernardo Bonanni, Irene Feroce, Mariarosaria Calvello, Aliana Guerrieri Gonzaga, Monica Marabelli, Davide Bondavalli and the personnel of the Cogentech Cancer Genetic Test Laboratory. The MCCS was made possible by the contribution of many people, including the original investigators, the teams that recruited the participants and continue working on follow-up, and the many thousands of Melbourne residents who continue to participate in the study. MYBRCA thanks study participants and research staff (particularly Patsy Ng, Nurhidayu Hassan, Yoon Sook-Yee, Daphne Lee, Lee Sheau Yee, Phuah Sze Yee and Norhashimah Hassan) for their contributions and commitment to this study. The following are NBCS Collaborators: Kristine K. Sahlberg (PhD), Anne-Lise Børresen-Dale (Prof. Em.), Lars Ottestad (MD), Rolf Kåresen (Prof. Em.) Dr. Ellen Schlichting (MD), Marit Muri Holmen (MD), Toril Sauer (MD), Vilde Haakensen (MD), Olav Engebråten (MD), Bjørn Naume (MD), Alexander Fosså (MD), Cecile E. Kiserud (MD), Kristin V. Reinertsen (MD), Åslaug Helland (MD), Margit Riis (MD), Jürgen Geisler (MD), OSBREAC and Grethe I. Grenaker Alnæs (MSc). NBHS and SBCGS thank study participants and research staff for their contributions and commitment to the studies. For NHS and NHS2 the study protocol was approved by the institutional review boards of the Brigham and Women's Hospital and Harvard T.H. Chan School of Public Health, and those of participating registries as required. We would like to thank the participants and staff of the NHS and NHS2 for their valuable contributions as well as the following state cancer registries for their help: AL, AZ, AR, CA, CO, CT, DE, FL, GA, ID, IL, IN, IA, KY, LA, ME, MD, MA, MI, NE, NH, NJ, NY, NC, ND, OH, OK, OR, PA, RI, SC, TN, TX, VA, WA, WY. The authors assume full responsibility for analyses and interpretation of these data. OBCS thanks Arja Jukkola-Vuorinen, Mervi Grip, Saila Kauppila, Meeri Otsukka, Leena Keskitalo and Kari Mononen for their contributions to this study. The OFBCR thanks Teresa Selander, Nayana Weerasooriya and Steve Gallinger. ORIGO thanks E. Krol-Warmerdam, and J. Blom for patient accrual, administering questionnaires, and managing clinical information. The LUMC survival data were retrieved from the Leiden hospital-based cancer registry system (ONCDOC) with the help of Dr. J. Molenaar. PBCS thanks Louise Brinton, Mark Sherman, Neonila Szeszenia-Dabrowska, Beata Peplonska, Witold Zatonski, Pei Chao, Michael Stagner. The ethical approval for the POSH study is MREC /00/6/69, UKCRN ID: 1137. We thank the staff in the Experimental Cancer Medicine Centre (ECMC) supported Faculty of Medicine Tissue Bank and the Faculty of Medicine DNA Banking resource. The authors wish to acknowledge the roles of the Breast Cancer Now Tissue Bank in collecting and making available the samples and/or data, and the patients who have generously donated their tissues and shared their data to be used in the generation of this publication. The RBCS thanks Jannet Blom, Saskia Pelders, Wendy J.C. Prager–van der Smissen, and the Erasmus MC Family Cancer Clinic. SBCS thanks Sue Higham, Helen Cramp, Dan Connley, Ian Brock, Sabapathy Balasubramanian and Malcolm W.R. Reed. We thank the SEARCH and EPIC teams. SGBCC thanks the participants and all research coordinators for their excellent help with recruitment, data and sample collection. SKKDKFZS thanks all study participants, clinicians, family doctors, researchers and technicians for their contributions and commitment to this study. UBCS thanks all study participants as well as the ascertainment, laboratory, analytics and informatics teams at Huntsman Cancer Institute and Intermountain Healthcare for their important contributions to this study. UCIBCS thanks Irene Masunaka.

### Supplementary Tables

Supplementary Tables 1-2, 4-5, 7, 9-10, 13

**Supplementary Table 1: Summary of numbers of cases and controls used in the analysis, as well as median age of participants.** Family history was a selection criterion for case inclusion in BRIDGES. The variable median age for cases is the median diagnosed age and for controls is the median age at the first assessment centre visit (UK Biobank) or observation (BRIDGES).

| | Female cases | Female controls | Male cases | Male controls |
|---|---|---|---|---|
| **Total** | | | | |
| **UK Biobank** | **8001** | **92534** | **38** | **82364** |
| No family history | 6495 | 82337 | 32 | 73785 |
| family history | 1506 | 10197 | 6 | 8579 |
| *median age* | *60* | *58* | *62* | *58* |
| **BRIDGES** | **51494** | **43884** | | |
| No family history | 32497 | 21496 | | |
| family history | 11994 | 3328 | | |
| family history not available | 7003 | 19060 | | |
| *median age* | *54* | *58* | | |

**Supplementary Table 2: Association results for the burden of rare variants in 5' UTR regions and breast cancer.** All regions associated at P<0.001 are listed. Z-scores are from testing H0 of no association. P-values are unadjusted for multiple testing.

| chr | UTR start | UTR end | Gene | Controls* | | Cases* | | Odds Ratio | Z-score | P-value |
|---|---|---|---|---|---|---|---|---|---|---|
| | | | | Non-Carriers | Carriers | Non-Carriers | Carriers | | | |
| 17 | 32486993 | 32487620 | *CDK5R1* | 91680 | 854 | 7878 | 123 | 1.48 (1.27, 1.73) | 4.92 | 8.49E-07 |
| 19 | 17420047 | 17420135 | *MVB12A* | 92440 | 94 | 7980 | 21 | 2.23 (1.5, 3.31) | 3.99 | 6.58E-05 |
| 6 | 152628332 | 152637362 | *SYNE1* | 92113 | 421 | 7947 | 54 | 1.55 (1.25, 1.93) | 3.95 | 7.95E-05 |
| 6 | 32164595 | 32166165 | *EGFL8* | 92503 | 31 | 7992 | 9 | 3.3 (1.8, 6.06) | 3.85 | 0.000118 |
| 17 | 45262017 | 45262094 | *SPATA32* | 92509 | 25 | 7993 | 8 | 3.5 (1.82, 6.76) | 3.74 | 0.000182 |
| 19 | 37594876 | 37598447 | *ZNF540* | 92394 | 140 | 7999 | 2 | 0.216 (0.0959, 0.484) | -3.71 | 0.000204 |
| 6 | 53041237 | 53061824 | *CILK1* | 92292 | 242 | 7966 | 35 | 1.67 (1.26, 2.21) | 3.61 | 0.000306 |
| 2 | 233060342 | 233060478 | *INPP5D* | 92461 | 73 | 7988 | 13 | 2.29 (1.44, 3.63) | 3.51 | 0.000447 |
| 2 | 263985 | 264032 | *SH3YL1* | 92491 | 43 | 7991 | 10 | 2.68 (1.54, 4.66) | 3.48 | 0.000497 |
| 20 | 62219971 | 62220278 | *HRH3* | 92205 | 329 | 7960 | 41 | 1.55 (1.21, 1.99) | 3.48 | 0.000498 |
| 7 | 151205455 | 151205496 | *IQCA1L* | 92463 | 71 | 7986 | 15 | 2.23 (1.42, 3.51) | 3.46 | 0.000548 |
| 10 | 69573044 | 69573422 | *NEUROG3* | 92353 | 181 | 7972 | 29 | 1.74 (1.27, 2.4) | 3.42 | 0.000617 |
| 11 | 68039025 | 68041271 | *TCIRG1* | 92431 | 103 | 7985 | 16 | 2.03 (1.35, 3.05) | 3.40 | 0.000662 |
| 6 | 127331029 | 127343609 | *ECHDC1* | 92348 | 186 | 7975 | 26 | 1.72 (1.26, 2.36) | 3.38 | 0.000721 |
| 1 | 154983344 | 154983694 | *FLAD1* | 92266 | 268 | 7957 | 44 | 1.58 (1.21, 2.07) | 3.37 | 0.000744 |
| 6 | 4135561 | 4135575 | *ECI2* | 92532 | 2 | 7997 | 4 | 9.37 (2.55, 34.4) | 3.37 | 0.000746 |
| 3 | 184174855 | 184176993 | *AP2M1* | 92292 | 242 | 7962 | 39 | 1.62 (1.22, 2.14) | 3.36 | 0.000764 |
| 22 | 50244460 | 50245023 | *TUBGCP6* | 92027 | 507 | 7946 | 55 | 1.43 (1.16, 1.76) | 3.35 | 0.000805 |
| 23 | 132127845 | 132128020 | *FRMD7* | 92399 | 135 | 7976 | 25 | 1.9 (1.3, 2.77) | 3.33 | 0.000856 |
| 7 | 141551410 | 141555466 | *AGK* | 92468 | 66 | 7987 | 14 | 2.24 (1.39, 3.59) | 3.33 | 0.000856 |
| 14 | 49620799 | 49621268 | *MGAT2* | 92083 | 451 | 7981 | 20 | 0.59 (0.431, 0.806) | -3.31 | 0.000942 |
| 18 | 9334773 | 9337229 | *TWSG1* | 92381 | 153 | 7980 | 21 | 1.78 (1.26, 2.51) | 3.30 | 0.000948 |
| 16 | 3112586 | 3113430 | *ZNF205* | 92492 | 42 | 7989 | 12 | 2.54 (1.46, 4.43) | 3.29 | 0.000997 |

*The counts are for female cases and controls only, but odds-ratios and results are from incorporating males and family history data - see methods

**Supplementary Table 4: P-values from SKAT-O and burden tests for rare variants in 5' UTR regions of the 5 known breast cancer risk genes.** P-values are from testing H0 of no association. P-values are unadjusted for multiple testing.

| chr | UTR start | UTR end | Gene | No. variants | Controls* | | Cases* | | Test | OR | P-value |
|---|---|---|---|---|---|---|---|---|---|---|---|
| | | | | | Non-Carriers | Carriers | Non-Carriers | Carriers | | | |
| 11 | 108223067 | 108227624 | *ATM* | 29 | 92281 | 253 | 7984 | 17 | SKATOrobust | NA | 0.46 |
| | | | | | | | | | Burden | 0.788 (0.545, 1.14) | 0.21 |
| 17 | 43124097 | 43125364 | *BRCA1* | 26 | 92357 | 177 | 7986 | 15 | SKATOrobust | NA | 1.00 |
| | | | | | | | | | Burden | 0.946 (0.628, 1.42) | 0.79 |
| 13 | 32315508 | 32316460 | *BRCA2* | 52 | 92379 | 155 | 7987 | 14 | SKATOrobust | NA | 1.00 |
| | | | | | | | | | Burden | 1.03 (0.675, 1.57) | 0.89 |
| 22 | 28734722 | 28741820 | *CHEK2* | 19 | 92462 | 72 | 7994 | 7 | SKATOrobust | NA | 0.42 |
| | | | | | | | | | Burden | 1.13 (0.618, 2.05) | 0.70 |
| 16 | 23641158 | 23641310 | *PALB2* | 23 | 92410 | 124 | 7992 | 9 | SKATOrobust | NA | 0.78 |
| | | | | | | | | | Burden | 0.88 (0.537, 1.44) | 0.61 |

*The counts are for female cases and controls only, but odds-ratios and results are from incorporating males and family history data - see methods

**Supplementary Table 5: Burden and SKAT-O tests for groups of variants in the CDK5R1 5' UTR region.**

| | N variants | Burden | | SKAT-O robust (females) |
|---|---|---|---|---|
| | | OR (LB, UB) | p-value | p-value |
| **All UTR variants** | 105 | 1.48 (1.27, 1.73) | $8.48 \times 10^{-7}$ | $7.19 \times 10^{-7}$ |
| **UTR variants,** <32487160 | 69 | 1.46 (1.23, 1.73) | $1.65 \times 10^{-5}$ | $1.94 \times 10^{-6}$ |
| **UTR variants,** >32487483 | 36 | 1.63 (1.14, 2.34) | 0.0081 | 0.241 |
| **Intron variants** | 60 | 1.28 (0.96, 1.73) | 0.097 | 0.115 |
| **All variants** | 168 | 1.43 (1.25, 1.65) | $3.43 \times 10^{-7}$ | $6.23 \times 10^{-7}$ |

**Supplementary Table 7**: **Association results for the burden of rare variants in 3' UTR regions and breast cancer.** All regions associated at P<0.001 are listed. P-values are from testing H0 of no association. P-values are unadjusted for multiple testing.

| chr | UTR start | UTR end | Gene | Controls* | | Cases* | | Odds Ratio | Z-score | P-value |
|---|---|---|---|---|---|---|---|---|---|---|
| | | | | Non-Carriers | Carriers | Non-Carriers | Carriers | | | |
| 1 | 154697455 | 154707975 | *KCNN3* | 85659 | 6875 | 7315 | 686 | 1.14 (1.07, 1.22) | 4.08 | 4.56E-05 |
| 16 | 71859685 | 71860017 | *ZNF821* | 92356 | 178 | 7965 | 36 | 1.8 (1.32, 2.46) | 3.68 | 0.000232 |
| 21 | 43030102 | 43033931 | *PKNOX1* | 89688 | 2846 | 7804 | 197 | 0.816 (0.731, 0.911) | -3.62 | 0.000297 |
| 2 | 169528508 | 169529824 | *FASTKD1* | 91720 | 814 | 7950 | 51 | 0.67 (0.539, 0.834) | -3.58 | 0.000338 |
| 19 | 35746848 | 35747519 | *PSENEN* | 92160 | 374 | 7991 | 10 | 0.514 (0.358, 0.74) | -3.58 | 0.000343 |
| 19 | 7682821 | 7687703 | *TRAPPC5* | 88167 | 4367 | 7691 | 310 | 0.854 (0.782, 0.932) | -3.54 | 0.000403 |
| 14 | 100530042 | 100530303 | *WDR25* | 92285 | 249 | 7967 | 34 | 1.65 (1.25, 2.18) | 3.52 | 0.000437 |
| 6 | 125759803 | 125761269 | *HEY2* | 91740 | 794 | 7904 | 97 | 1.35 (1.14, 1.6) | 3.47 | 0.000530 |
| 3 | 15254353 | 15256085 | *SH3BP5* | 90802 | 1732 | 7880 | 121 | 0.779 (0.676, 0.898) | -3.45 | 0.000550 |
| 12 | 49762897 | 49764934 | *TMBIM6* | 90665 | 1869 | 7878 | 123 | 0.785 (0.685, 0.901) | -3.45 | 0.000558 |
| 20 | 23495915 | 23496010 | *CST8* | 92510 | 24 | 7998 | 3 | 3.52 (1.72, 7.21) | 3.44 | 0.000575 |
| 11 | 62888694 | 62888860 | *SLC3A2* | 92438 | 96 | 7982 | 19 | 2.01 (1.34, 3.02) | 3.39 | 0.000701 |
| 3 | 112081085 | 112081269 | *TMPRSS7* | 92521 | 13 | 7995 | 6 | 4.25 (1.84, 9.82) | 3.38 | 0.000723 |
| 5 | 160347754 | 160349167 | *C1QTNF2* | 91244 | 1290 | 7916 | 85 | 0.751 (0.636, 0.888) | -3.36 | 0.000779 |
| 3 | 193274789 | 193275041 | *ATP13A5* | 92300 | 234 | 7968 | 33 | 1.63 (1.23, 2.18) | 3.35 | 0.000804 |
| 8 | 144930358 | 144930737 | *ZNF16* | 92331 | 203 | 7970 | 31 | 1.69 (1.24, 2.3) | 3.34 | 0.000824 |
| 10 | 97865000 | 97865547 | *CRTAC1* | 92132 | 402 | 7947 | 54 | 1.47 (1.17, 1.85) | 3.32 | 0.000893 |
| 23 | 105218929 | 105218963 | *TEX13A* | 92531 | 3 | 7999 | 2 | 15.4 (3.03, 78.6) | 3.29 | 0.000992 |

*The counts are for female cases and controls only, but odds-ratios and results are from incorporating males and family history data - see methods

**Supplementary Table 9: P-values from SKAT-O and burden tests for rare variants in 3' UTR regions of the 5 known breast cancer risk genes**. P-values are from testing H0 of no association. P-values are unadjusted for multiple testing.

| chr | UTR start | UTR end | Gene | No. variants | Controls* | | Cases* | | Test | OR | P-value |
|---|---|---|---|---|---|---|---|---|---|---|---|
| | | | | | Non-Carriers | Carriers | Non-Carriers | Carriers | | | |
| 11 | 108365509 | 108369102 | *ATM* | 539 | 88996 | 3538 | 7696 | 305 | SKATOrobust | | 0.850 |
| | | | | | | | | | Burden | 1.05 (0.957, 1.15) | 0.317 |
| 17 | 43044295 | 43045677 | *BRCA1* | 172 | 91888 | 646 | 7957 | 44 | SKATOrobust | | 0.116 |
| | | | | | | | | | Burden | 0.860 (0.688, 1.08) | 0.186 |
| 13 | 32398771 | 32400268 | *BRCA2* | 130 | 91879 | 655 | 7937 | 64 | SKATOrobust | | 0.583 |
| | | | | | | | | | Burden | 1.17 (0.962, 1.43) | 0.116 |
| 22 | 28687743 | 28687896 | *CHEK2* | 22 | 92345 | 189 | 7979 | 22 | SKATOrobust | | 0.288 |
| | | | | | | | | | Burden | 1.36 (0.961, 1.92) | 0.0825 |
| 16 | 23603165 | 23603458 | *PALB2* | 26 | 92457 | 77 | 7998 | 3 | SKATOrobust | | 0.297 |
| | | | | | | | | | Burden | 0.458 (0.200, 1.05) | 0.0657 |

*The counts are for female cases and controls only, but odds-ratios and results are from incorporating males and family history data - see methods

**Supplementary Table 10: Meta-analysis P-values from burden tests for rare variants in 3' and 5' UTR regions and breast cancer for 35 BRIDGES genes.** Regions with $P_{BM}$<0.1 are listed. The table is sorted by $P_{BM}$. P-values are from testing H0 of no association. P-values are unadjusted for multiple testing.

| chr | UTR start | UTR end | Gene | UTR number | Dataset | Controls* | | Cases* | | OR | Z-score | P-value | Meta-analysed Z-score | Meta-analysed P-value |
|---|---|---|---|---|---|---|---|---|---|---|---|---|---|---|
| | | | | | | Non-Carriers | Carriers | Non-Carriers | Carriers | | | | | |
| 2 | 214809570 | 214809683 | *BARD1* | 5 | BRIDGES | 43880 | 4 | 51492 | 2 | 0.430 (0.0916, 2.02) | -1.07 | 0.286 | -2.67 | 0.00762 |
| | | | | | UKB | 92476 | 58 | 7999 | 2 | 0.190 (0.0538, 0.673) | -2.57 | 0.0101 | | |
| 22 | 28687743 | 28687896 | *CHEK2* | 3 | BRIDGES | 43870 | 14 | 51457 | 37 | 1.573 (0.928, 2.66) | 1.68 | 0.0920 | 2.38 | 0.0175 |
| | | | | | UKB | 92345 | 189 | 7979 | 22 | 1.36 (0.961, 1.92) | 1.74 | 0.0825 | | |
| 7 | 105532201 | 105532315 | *RINT1* | 5 | BRIDGES | 43839 | 45 | 51424 | 70 | 1.24 (0.887, 1.73) | 1.26 | 0.209 | 2.37 | 0.0179 |
| | | | | | UKB | 92435 | 99 | 7988 | 13 | 1.65 (1.07, 2.54) | 2.25 | 0.0244 | | |
| 17 | 43124097 | 43125364 | *BRCA1* | 5 | BRIDGES | 43772 | 112 | 51324 | 170 | 1.30 (1.05, 1.61) | 2.37 | 0.0177 | 1.97 | 0.0485 |
| | | | | | UKB | 92357 | 177 | 7986 | 15 | 0.946 (0.628, 1.43) | -0.27 | 0.789 | | |
| 17 | 31094977 | 31095309 | *NF1* | 5 | BRIDGES | 43790 | 94 | 51393 | 101 | 0.768 (0.594, 0.994) | -2.01 | 0.0447 | -1.92 | 0.0553 |
| | | | | | UKB | 92287 | 247 | 7981 | 20 | 0.914 (0.644, 1.30) | -0.50 | 0.615 | | |
| 16 | 23603165 | 23603458 | *PALB2* | 3 | BRIDGES | 43882 | 2 | 51494 | 0 | 0.00 (0.00, Inf) | -0.02 | 0.987 | -1.84 | 0.0657 |
| | | | | | UKB | 92457 | 77 | 7998 | 3 | 0.458 (0.200, 1.05) | -1.84 | 0.0657 | | |

*The counts are for female cases and controls only, but odds-ratios and results are from incorporating males and family history data - see methods

**Supplementary Table 13: P-values from SKAT-O and burden tests for rare variants in promoter regions of the 5 known breast cancer risk genes.** P-values are from testing H0 of no association. P-values are unadjusted for multiple testing.

| chr | promoter start | promoter end | Regulatory stable ID | Gene | No. variants | Controls* | | Cases* | | Test | OR | P-value |
|---|---|---|---|---|---|---|---|---|---|---|---|---|
| | | | | | | Non-Carriers | Carriers | Non-Carriers | Carriers | | | |
| 11 | 108220000 | 108227401 | ENSR00000044515 | *ATM* | 1195 | 86907 | 5627 | 7489 | 512 | SKATOrobust | | 0.516 |
| | | | | | | | | | | Burden | 1.03 (0.959, 1.11) | 0.412 |
| 17 | 43123800 | 43127201 | ENSR00000094515 | *BRCA1* | 555 | 89493 | 3041 | 7748 | 253 | SKATOrobust | | 0.666 |
| | | | | | | | | | | Burden | 0.985 (0.892, 1.09) | 0.763 |
| 13 | 32314600 | 32317001 | ENSR00000060894 | *BRCA2* | 303 | 90954 | 1580 | 7844 | 157 | SKATOrobust | | 0.139 |
| | | | | | | | | | | Burden | 1.15 (1.01, 1.30) | 0.0392 |
| 22 | 28740400 | 28743800 | ENSR00000144944 | *CHEK2* | 479 | 90235 | 2299 | 7796 | 205 | SKATOrobust | | 0.723 |
| | | | | | | | | | | Burden | 1.01 (0.903, 1.13) | 0.857 |
| 16 | 23639600 | 23643200 | ENSR00000084571 | *PALB2* | 520 | 89811 | 2723 | 7764 | 237 | SKATOrobust | | 1.00 |
| | | | | | | | | | | Burden | 1.07 (0.971, 1.19) | 0.164 |

*The counts are for female cases and controls only, but odds-ratios and results are from incorporating males and family history data - see methods

Excel Supplementary Table legends

**Supplementary Table 3: P-values from SKAT-O and burden tests for rare variants in 5' UTR regions and breast cancer, for regions with $P_B$<0.05.** Regions with $P_{min}$<5x10-5 are listed, where $P_{min}$=min($P_B$, $P_S$). The table is sorted by $P_{min}$. P-values are from testing H0 of no association. P-values are unadjusted for multiple testing.

**Supplementary Table 6: UTR variants in the 5' UTR CDK5R1 region.** Variants are sorted by carrier frequency.

**Supplementary Table 8: P-values from SKAT-O and burden tests for rare variants in 3' UTR regions and breast cancer, for regions with $P_B$<0.05.** Regions with $P_{min}$<5x10-5 are listed, where $P_{min}$=min($P_B$, $P_S$). The table is sorted by $P_{min}$. P-values are from testing H0 of no association. P-values are unadjusted for multiple testing.

**Supplementary Table 11: Association results for the burden of rare variants in promoter regions and breast cancer.** All regions associated at P<0.001 are listed. Z-scores are from testing H0 of no association. P-values are unadjusted for multiple testing.

**Supplementary Table 12: P-values from SKAT-O and burden tests for rare variants in promoter regions and breast cancer for regions with $P_B$<0.05** Regions with $P_{min}$<5x10-5 are listed, where $P_{min}$=min($P_B$, $P_S$). The table is sorted by $P_{min}$. P-values are from testing H0 of no association. P-values are unadjusted for multiple testing.
